# Accounting for bias due to outcome data missing not at random: comparison and illustration of two approaches to probabilistic bias analysis: a simulation study

**DOI:** 10.1101/2024.03.24.24304792

**Authors:** Emily Kawabata, Daniel Major-Smith, Gemma L Clayton, Chin Yang Shapland, Tim P Morris, Alice R Carter, Alba Fernández-Sanlés, Maria Carolina Borges, Kate Tilling, Gareth J Griffith, Louise AC Millard, George Davey Smith, Deborah A Lawlor, Rachael A Hughes

**Affiliations:** MRC Integrative Epidemiology Unit, University of Bristol, Bristol, UK; Population Health Sciences, Bristol Medical School, University of Bristol, Bristol, UK; MRC Clinical Trials Unit at UCL, London, UK; MRC Unit for Lifelong Health and Ageing at University College London, London, UK

**Author notes:** These authors contributed equally to this work (Emily Kawabata, Daniel Major-Smith, Gemma L Clayton).

**Keywords:** Bayesian bias analysis, inverse probability weighting, missing not at random, Monte Carlo bias analysis, multiple imputation, probabilistic bias analysis, sensitivity analysis; UK Biobank

## Abstract

**Background:** Bias from data missing not at random (MNAR) is a persistent concern in health-related research. A bias analysis quantitatively assesses how conclusions change under different assumptions about missingness using bias parameters which govern the magnitude and direction of the bias. Probabilistic bias analysis specifies a prior distribution for these parameters, explicitly incorporating available information and uncertainty about their true values. A Bayesian approach combines the prior distribution with the data’s likelihood function whilst a Monte Carlo approach samples the bias parameters directly from the prior distribution. No study has compared a Monte Carlo approach to a fully Bayesian approach in the context of a bias analysis to MNAR missingness.

**Methods:** We propose an accessible Monte Carlo probabilistic bias analysis which uses a well-known imputation method. We designed a simulation study based on a motivating example from the UK Biobank study, where a large proportion of the outcome was missing and missingness was suspected to be MNAR. We compared the performance of our Monte Carlo probabilistic bias analysis to a principled Bayesian probabilistic bias analysis, complete case analysis (CCA) and missing at random implementations of inverse probability weighting (IPW) and multiple imputation (MI).

**Results:** Estimates of CCA, IPW and MI were substantially biased, with 95% confidence interval coverages of 7–64%. Including auxiliary variables (i.e., variables not included in the substantive analysis which are predictive of missingness and the missing data) in MI’s imputation model amplified the bias due to assuming missing at random. With reasonably accurate and precise information about the bias parameter, the Monte Carlo probabilistic bias analysis performed as well as the fully Bayesian approach. However, when very limited information was provided about the bias parameter, only the Bayesian approach was able to eliminate most of the bias due to MNAR whilst the Monte Carlo approach performed no better than the CCA, IPW and MI.

**Conclusion:** Our proposed Monte Carlo probabilistic bias analysis approach is easy to implement in standard software and is a viable alternative to a Bayesian approach. We caution careful consideration of choice of auxiliary variables when applying imputation where data may be MNAR.

## Introduction

The main aim of many epidemiology studies is to estimate the causal effect of an exposure on an outcome (here onward, shortened to *exposure effect*). Inference about the exposure effect may be invalid when the sample included in the analysis is not a representative (random) sample of the target population. For example, this could be due to selection out of the study due to missing data. The choice of method for dealing with missing data partly depends on the mechanism causing the data to be missing (called *missingness mechanisms*). These mechanisms are commonly classified as missing completely at random (probability of missingness is independent of the observed and missing data), missing at random (MAR; probability of missingness is independent of the missing data given the observed) and missing not at random (MNAR; probability of missingness depends on the missing data even after conditioning on the observed data) (1). We focus on a MNAR missingness mechanism where the value of a variable directly affects its own probability of missingness (2). This type of MNAR missingness mechanism commonly occurs when collecting sensitive information (such as information on sexual health, financial matters, and substance-use behaviours) or when the health outcome under study affects participation. Routinely used missing data methods, such as multiple imputation (MI) and inverse probability weighting (IPW), assume data are MAR and may give biased results when the missingness mechanism is MNAR.

Information about the missingness mechanism may be available from ancillary data such as instruments for missingness (3), record-linkage data (4,5), and responsiveness data (6). In the absence of such information, the analyst cannot distinguish between MAR and MNAR missingness mechanisms based on the observed data only (7). Instead, the analyst must base their decision on expert knowledge or available literature. When MNAR missingness is suspected, a bias analysis is recommended to quantify the potential impact of MNAR missingness on their study conclusions (8,9,10).

A bias analysis for MNAR missingness (here onward, shortened to *bias analysis*) requires a model (known as a bias model) for the data and missingness mechanism. Two commonly used approaches are selection models and pattern-mixture models (8). Without any loss of generality, in the context of an outcome MNAR the selection model usually consists of a model for the substantive analysis of interest (fitted to participants with observed and missing outcome) and a model for the missingness mechanism that characterizes how missingness depends on the outcome. In contrast, the pattern-mixture model describes how the distribution of the outcome depends on missingness and may consist of a model for the substantive analysis that differs between participants with observed and missing outcome. Both types of model can be fitted using maximum likelihood, within a Bayesian framework or using multiple imputation (8).

For both the selection and pattern-mixture models, under MNAR the observed data does not provide any information about the parameters governing the dependency between the outcome and missingness (known as bias or sensitivity parameters). Scharfstein et al argued that the bias parameters are more plausibly a priori independent of the substantive analysis parameters in a selection model than in a pattern-mixture model (11). However, others have argued that the bias parameters of a pattern-mixture model are usually easier to interpret than those of a selection model (12,13). For both models, setting the bias parameters to prespecified values enables estimation of the remaining parameters of the model and provides an estimate of the exposure effect adjusted for bias due to MNAR (here onward, called the *bias-adjusted exposure effect estimate*). Changing the values of these bias parameters allows estimation of the bias-adjusted exposure effect under different assumptions about the missingness mechanism.

A bias analysis can be implemented as a deterministic or probabilistic bias analysis (9). In a deterministic bias analysis, a range of values is specified for all bias parameters and then for each plausible combination of values, the bias model is estimated by fixing the bias parameters to these values. This approach provides the analyst with information about the range of possible estimates for the exposure effect but does not indicate which of these estimates are most likely to occur, making interpretation of the results challenging (9). Alternatively, a probabilistic bias analysis specifies a prior probability distribution for the bias parameters which explicitly incorporates the analyst’s assumptions about plausible values and the combinations of values most likely to occur. The probabilistic bias analysis generates a distribution of bias-adjusted exposure effect estimates which is then summarised as a point estimate (e.g., the median as a measure of central tendency) and a 95% interval estimate (e.g., 2.5^th^ and 97.5^th^ percentiles as limits of the interval) that accounts for the analyst’s uncertainty about the MNAR missingness mechanism in addition to the usual random sampling error.

A probabilistic bias analysis can be implemented as a fully Bayesian analysis where the prior distribution is combined with the likelihood function for the data and the resulting posterior distribution is then summarised (14). The alternative is a Monte Carlo approach which repeatedly samples the bias parameters directly from its prior distribution, uses these sampled values to estimate the bias-adjusted exposure effect, and then incorporates random sampling error to give a frequency distribution of bias-adjusted exposure effect estimates (14). Generally, the Monte Carlo approach is simpler to understand, quicker and easier to implement as it requires no Bayesian computation (9,15,16).

In the context of bias analysis to unmeasured confounding or misclassification, a small number of studies have compared the Monte Carlo approach to a fully Bayesian analysis (14,15,16,17,18,19). Along with some theoretical arguments, these studies indicate that the Monte Carlo approach is a good approximation of a fully Bayesian analysis provided the prior distribution for the bias parameters only specifies plausible values given the observed data (e.g., among healthy adults, sampled values for the mean difference in body mass index (BMI) between participants with observed and missing values always implies missing BMI values are greater than 18 kg/m^2^) (15,16,17,18). In the presence of implausible values for the bias parameters, the Monte Carlo approach can give interval estimates that are either too wide or too narrow (14,17). No study has compared a Monte Carlo approach to a fully Bayesian analysis in the context of a bias analysis to MNAR missingness.

Despite significant developments in bias analysis methods and software in the past decade, bias analyses are not routinely reported (20). This indicates a large gap between methodological development and practical application. In the current literature, there is limited guidance on implementing a probabilistic bias analysis to data MNAR. Recent exceptions for cross-sectional analyses include: (1) a pattern-mixture approach where draws from a prior distribution (of the bias parameters) are used to impute a categorical covariate under MNAR (20,21) and (2) a Bayesian implementation of a selection model for a partially observed continuous outcome (22). Additionally, in the context of selection bias due to non-random selection of participants into a study, Banack et al review and compare a Monte Carlo probabilistic bias analysis to an alternative approach that simulates the entire dataset under different assumptions about the selection bias (23) and Jayaweera et al conducted a Monte Carlo probabilistic bias analysis by inversely weighting participants based on their probability of inclusion (i.e., participating and remaining in the study combined) (24).

In this paper, we illustrate a Monte Carlo probabilistic bias analysis using a pattern-mixture version of the commonly used imputation method fully conditional specification (also known as chained equations) (25,26,27). Via a data example and simulations, we compare the performance of the proposed Monte Carlo probabilistic bias analysis to a fully Bayesian probabilistic bias analysis in a setting where a large proportion of the outcome is missing and missingness is suspected to be affected by the outcome’s own values. We begin with a brief description of principled missing data methods, including our proposed Monte Carlo probabilistic bias analysis and a fully Bayesian probabilistic bias analysis, illustrated using a hypothetical example. We then describe the simulation study and application of the approaches to a motivating case study. We conclude with a general discussion. R and Stata software code implementing the proposed Monte Carlo and the fully Bayesian probabilistic bias analyses is available from https://github.com/MRCIEU/COVIDITY_ProbQBA.

## Methods

### Hypothetical example

We want to estimate the effect of an exposure (or treatment) *X* on an outcome *Y* and we denote this exposure effect by *β*_*X*_. To estimate *β*_*X*_, our substantive analysis of interest is a generalised linear regression of *Y* on *X* adjusted for measured confounders *Z* and *W*

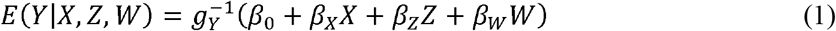

where 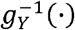 denotes the inverse link function (e.g., inverse of the logit function for logistic regression). We assume all confounders of the *Y − X* association are measured and without error, and in the absence of missing data that the substantive analysis would give unbiased results for *β*_*X*_. Outcome *Y* is observed in a small proportion of study participants. The study recorded data on auxiliary variables (i.e., variables not included in the substantive analysis) which are predictive of the missing values of *Y* and whether *Y* was observed or missing. In the full sample (participants with observed and missing values for *Y*), exposure *X* and some of the confounders and auxiliary variables were partially observed. Let *Z* and *W* denote the fully and partially observed confounders, respectively, and *A* and *D* denote the fully and partially observed auxiliary variables, respectively. To simplify the notation, and without loss of generality, we assume that *A* denotes a single variable, and similarly for *D, Z* and *W*. Binary variables *M*^*Y*^, *M*^*X*^, *M*^*W*^ and *M*^*D*^ denote the missingness indicators of *Y, X, W* and *D*, respectively (e.g., *M*^*Y*^ =1 when *Y* is missing and *M*^*Y*^ = 0 otherwise). The notation is summarised in Table 1.

**Table 1:**
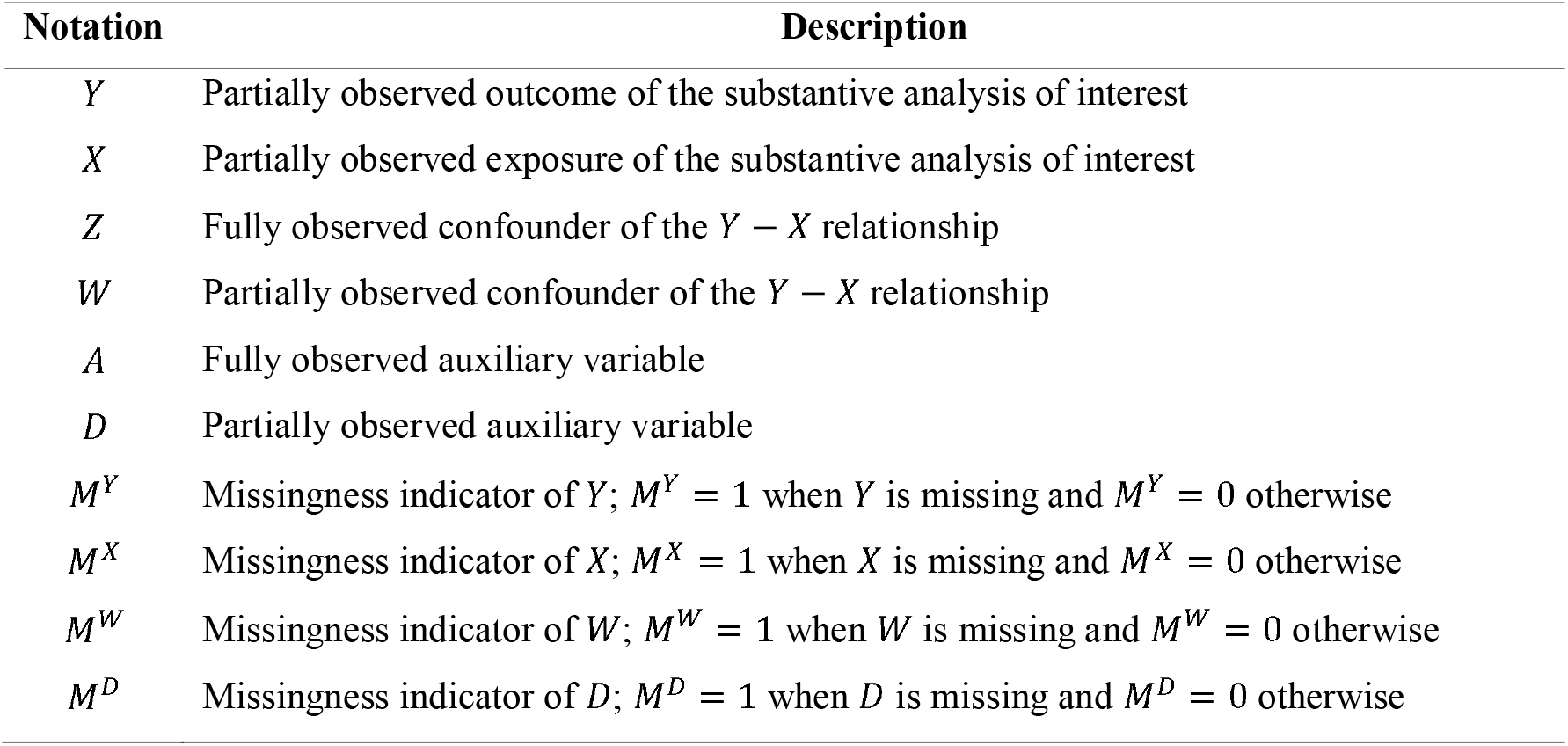
Description of the variables of the hypothetical example

| Notation | Description |
| --- | --- |
| $Y$ | Partially observed outcome of the substantive analysis of interest |
| $X$ | Partially observed exposure of the substantive analysis of interest |
| $Z$ | Fully observed confounder of the $Y - X$ relationship |
| $W$ | Partially observed confounder of the $Y - X$ relationship |
| $A$ | Fully observed auxiliary variable |
| $D$ | Partially observed auxiliary variable |
| $M^Y$ | Missingness indicator of $Y$; $M^Y = 1$ when $Y$ is missing and $M^Y = 0$ otherwise |
| $M^X$ | Missingness indicator of $X$; $M^X = 1$ when $X$ is missing and $M^X = 0$ otherwise |
| $M^W$ | Missingness indicator of $W$; $M^W = 1$ when $W$ is missing and $M^W = 0$ otherwise |
| $M^D$ | Missingness indicator of $D$; $M^D = 1$ when $D$ is missing and $M^D = 0$ otherwise |

Figure 1 depicts two missingness directed acyclic graphs (m-DAGs (28)) showing the relationships among the variables of our substantive analysis of interest (*Y, X, Z* and *W*), the auxiliary variables (*A* and *D*), and the missingness mechanisms of *Y* (depicted by blue edges), and of *X, W* and *D* (depicted by red edges). We note that exposure effect, *β*_*X*_, represents the total effect of *X* on *Y* (i.e., direct effect and indirect effect via auxiliaries A and D). We consider two scenarios, when *β*_*X*_ is not-null (figure 1a) and null (figure 1b). Note that *U*^*WZ*^ and *U*^*DA*^ denote unmeasured shared ancestors of *W* and *Z*, and *D* and *A*, respectively. Outcome *Y* is MNAR depending on fully observed auxiliary *A*, the missing values of *Y*, and the observed and missing values of exposure *X* and auxiliary *D*. We exclude the special case where the MNAR mechanism depends on *X* and *Y* independently (29). Variables *X, W* and *D* are MAR depending on fully observed confounder *Z* and auxiliary *A*; hence this MAR mechanism applies across all missing data patterns of *Y, X, W*, and *D*. In our example (but not depicted by the m-DAGs), the majority of participants with partially observed data are missing *Y* and a small proportion is missing data on *X, W* or *D* (with *Y* observed or missing).

**Figure 1:**
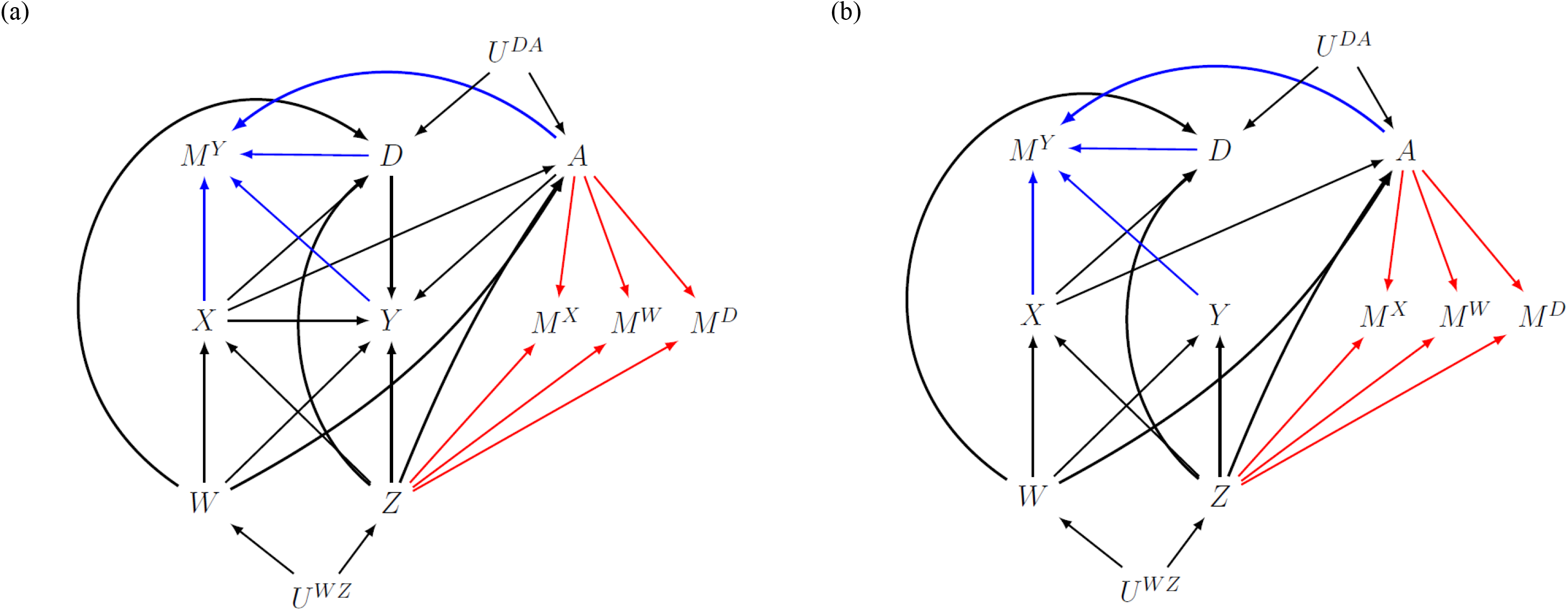
Missingness directed acyclic graphs (m-DAGs) of the scenario investigated by the simulation study when the exposure effect, *β*_*X*_, is (a) not-null and (b) null. Black edges depict the relationships in the fully observed data, and the blue and red edges depict the missingness mechanisms of the outcome and baseline variables (exposure, confounders, and auxiliary variables), respectively.

### Ignorable missing data methods

Ignorable missing data methods do not require a model for the joint distribution of the full data (observed and missing data) and the missingness mechanism (8). Popular ignorable methods include complete case analysis (CCA), and standard implementations of MI and IPW which assume MAR (hereafter referred to as MI and IPW, respectively) (7).

CCA fits the substantive analysis to participants with fully observed data on *Y, X, Z*, and *W* (known as complete cases). When the substantive analysis is a generalised linear regression, a CCA gives unbiased results when the probability of being a complete case does not depend on *Y* after conditioning on *X, z*, and *W* which includes some MNAR missingness mechanisms such as *X* MNAR depending on itself (7). Some additional exceptions apply for logistic regression as described in (7,29).

IPW is a type of weighted CCA where complete cases are weighted by the inverse of their probability of being a complete case (30). It is a two-stage process where the weights are estimated at stage 1 using a model which we shall refer to as the “weighting model”. At stage 2, these weights are used to fit the substantive analysis as a weighted CCA, where the standard errors are typically estimated using a sandwich estimator with the weights treated as known (30). Counterintuitively, ignoring uncertainty in the weights causes over-estimation of the standard errors (31). Typically, the weighting model is a logistic regression with a binary indicator variable for a complete case (1 if a complete case and 0 otherwise) as the dependent variable and the independent variables are predictors of being a complete case. These predictors can include variables of the substantive analysis or auxiliary variables. The validity of IPW requires completeness of a case to be independent of *Y, X, Z*, and *W* given these predictors and for the weighting model to be correctly specified (30).

It is challenging to implement IPW when the predictors of being a complete case are partially observed and the pattern of missingness is non-monotone (i.e., impossible to order the variables such that for any participant, if the *j*^*th*^ variable is observed then all of the previous *j −* 1 variables in the ordering are also observed) (30,32). To avoid this complication, IPW is mainly implemented using fully observed predictors. Another problem is inflated standard errors due to unstable weights (i.e., very large weights for some participants), which can arise when there are strong predictors of missingness. Using stabilised weights can curtail this inflation (30).

MI uses an imputation model to randomly sample values of the missing data (known as imputations) from their predictive distribution given the observed data (8). The missing values are replaced by the sampled values to form an imputed dataset. Multiple imputed datasets are generated, and the analysis model is fitted to each of these in turn. The results from the multiple imputed datasets are combined using Rubin’s rules to generate an average point estimate and a standard error that reflects the uncertainty about the missing values (8). The imputation model should include all variables of the substantive analysis. The validity of MI requires the data to be MAR given the variables of the imputation model and for the imputation model to be correctly specified. Auxiliary variables can be added to the imputation model to improve the plausibility of the MAR assumption or to improve precision. However, there are instances in which including auxiliary variables can amplify existing bias due to MNAR such as when auxiliary variables are strong predictors of the missing data (33) or are only predictive of missingness (34).

For our hypothetical example, CCA and MAR implementations of IPW and MI are expected to give a biased estimate for *β*_*X*_ in both the null and not-null scenarios. Since missingness of *Y* depends jointly on *X* and *Y*, CCA is an invalid approach even when the substantive analysis is a logistic regression (29). For IPW and MI, the MAR assumption is not achievable, regardless of the variables included in the weighting and imputation models, since missingness of *Y* depends directly on *Y* (i.e., path *Y →M*^*Y*^). However, for the not-null scenario, including A and D in the weighting and imputation models could eliminate some of the bias since missingness also depends on *Y* indirectly via *A* and *D* (e.g., path *Y ← A → M*^*Y*^ ).

We next describe two non-ignorable missing data methods, a fully Bayesian probabilistic bias analysis using a selection model (here onward, called *Bayesian SM*) and a Monte Carlo probabilistic bias analysis using a pattern-mixture model (here onward, called *Monte Carlo NARFCS*). The bias models of Bayesian SM and Monte Carlo NARFCS consist of a collection of generalised linear regressions. For simplicity and without loss of generality, we describe Bayesian SM and Monte Carlo NARFCS with respect to continuous variable *X* and binary variables *Y, W, A* and *D*, whilst *Z* is left unspecified and, by definition, missingness indicators *M*^*Y*^,*M*^*X*^, *M*^*W*^ and *M*^*D*^ are binary.

### Bayesian SM

#### Bias model specified as a selection model

We use the sequential modelling approach (35,36,37,38) to jointly model the substantive analysis, the MNAR missingness mechanism for *Y*, and models to “impute” (or estimate) the missing values of *X, W* and *D*. The sequential modelling approach factorises a joint distribution into a sequence of simpler univariate distributions, where each univariate distribution is modelled using an appropriate regression model (e.g., linear regression for continuous variables and logistic regression for binary variables). We specify the following regression models for the joint distribution of *W, X, Y, A, D, M*^*Y*^|*Z* :

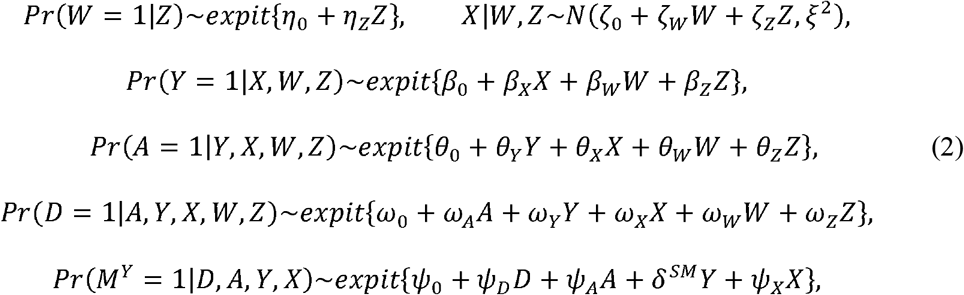

where *expit{k} = exp{k}/*1+ *exp{k}, M*^*Y*^ is assumed to be independent of *W* and *Z* given *D, A, Y* and *X* (as specified in figure 1), and *δ*^*SM*^ is the bias parameter representing the difference in the log-odds of observing *Y* between those with *Y=* 1 and *Y=* 0, conditional on *D, A, Y* and *X*. Let Ψ^*SM*^ denote the set of all estimable parameters of model (2) (i.e., all except *δ*^*SM*^), noting Ψ^*SM*^ includes exposure effect *β*_*X*_.

Theoretically, different orderings of these regression models may result in different joint distributions (39). We specified this ordering because it includes: the substantive analysis, a model for the MNAR missingness mechanism of *Y*, and incorporates auxiliary variables *A* and *D* without altering the substantive analysis. This ordering is compatible with a selection model framework. The ordering of the remaining models can be with respect to the amount of missing data (i.e., starting with the model for the variable with the least amount of missing data). Note that previous studies have reported that the fully Bayesian implementation of the sequential modelling approach appears robust to the ordering of the models (40,41) but the ordering may affect computational time (35).

#### Prior probability distributions

We assign independent prior distributions for all parameters. Following standard practice, we assign a normal distribution for each coefficient of the regression models and an inverse gamma distribution for the variance parameter of a linear regression (42). For *δ*^*SM*^ we assign Normal distribution *δ*^*SM*^ *∼N(μ*^*SM*^, *σ*^*SM*^) where values for mean *μ*^*SM*^ and variance *σ*^*SM*^ are chosen based on external information such as results from a published study. For all remaining parameters, Ψ^*SM*^, we assign vague priors; namely, *N*(0,100) for the coefficients and Inv-Gamma(0.01,0.01) for the variances.

#### Bayesian implementation

In the Bayesian framework, Bayes’ theorem is applied to combine the prior distributions for the bias model parameters with the likelihood function for the data to obtain the joint posterior distribution of (*δ*^*SM*^, Ψ^*SM*^). Therefore, application of Bayes’ theorem may rule out certain values of *δ*^*SM*^ because they are incompatible with the data (14). From the joint posterior distribution of (*δ*^*SM*^, Ψ^*SM*^), we can derive the conditional posterior distribution of a single parameter, such as *β*_*X*_.

The Bayesian framework views the missing data of *Y, X, W*, and *D* and parameters *δ*^*SM*^ and Ψ^*SM*^ as unknown quantities to be estimated. Since direct sampling from the joint distribution of these unknown quantities is difficult, we fit the selection model using Markov chain Monte Carlo (MCMC) estimation, specifically Gibbs sampling implemented by JAGS (version 4.3.0) (43,44,45) using R package *jagsUI* (version 1.5.2) (46).

### Monte Carlo NARFCS

#### Bias model specified as a pattern-mixture model

We use the Not-At-Random Fully Conditional Specification (NARFCS) approach (27) which is an MNAR extension of the MAR imputation method known as Fully Conditional Specification (FCS) (47). Like FCS, NARFCS imputes each variable under a separate univariate regression model (of type appropriate to the variable being imputed) and updates the missing data for each variable in turn using an iterative algorithm which we shall call the FCS algorithm (48,49). Note that the univariate distributions implied by these regression models may not be consistent with the same joint distribution and different orderings of these regression models within the FCS algorithm could lead to sampling from different joint distributions (25,50). The order in which the partially observed variables are updated within the FCS algorithm is typically determined by the amount of missing data (i.e., from the smallest to the greatest) (26).

We specify the following regression models for our NARFCS bias model:

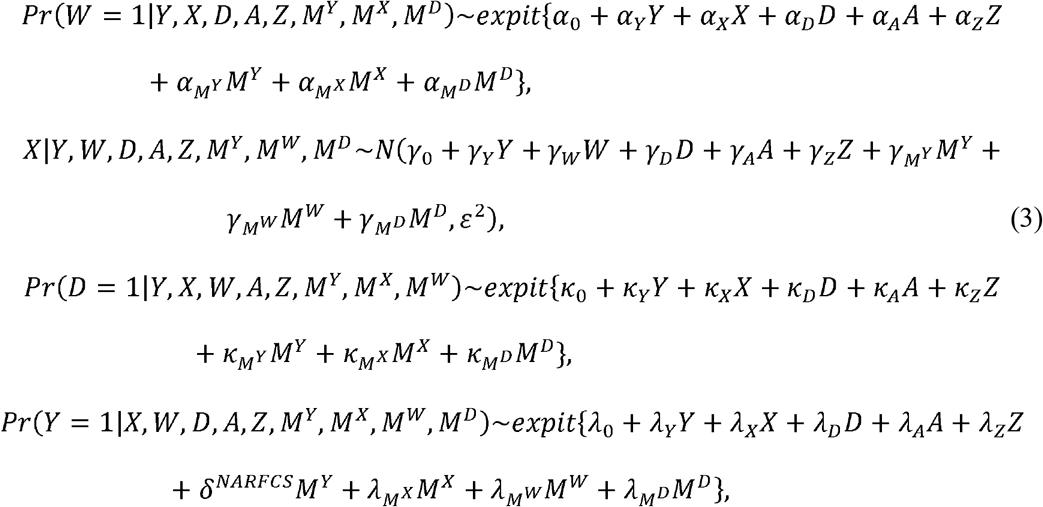

where *δ*^*NARFCS*^ is the bias parameter, representing the difference in the log-odds of *Y*= 1 between those with observed and missing values of *Y*. Let Ψ^*NARFCS*^ denote the set of all estimable parameters of model (3) (i.e., all except *δ*^*NARFCS*^ ), which does not include *β*_*X*_. Instead, an estimate of *β*_*X*_ is obtained by fitting the substantive analysis to the imputed data generated by the NARFCS bias model.

NARFCS differs from FCS in two ways which we shall illustrate using the regression model for *Y* in the bias model, (3), above. First, NARFCS includes missingness indicator *M*^*Y*^ as an independent variable in the regression model for *Y* in order to quantify how the distribution of *Y* differs between participants with observed and missing values of *Y*. Hence NARFCS belongs to the class of pattern-mixture models. Second, NARFCS includes the missingness indicators of the other partially observed variables, *M*^*X*^, *M*^*W*^,*M*^*D*^, as independent variables in the regression model for *Y* in order to maximise the amount of correlation between the variables captured by the model (27). Regarding the remaining regression models of (3), we note that the regression model for *X* omits *M*^*X*^ as an independent variable because we assume *X* is MAR given *A* and *Z*. Similarly, for the regression models of *W* and *D*.

#### Prior probability distributions

NARFCS does not assign a prior distribution for *δ*^*NARFCS*^ as it must be fixed to a prespecified value before applying the FCS algorithm (i.e., incorporated into the regression model for *Y* via a fixed offset term, *δ*^*NARFCS*^ *M*^*Y*^). For remaining parameters, Ψ^*NARFCS*^, NARFCS independently samples the parameters of each regression model from a posterior distribution (or an approximation) under a vague prior distribution (to ensure uncertainty from estimating the imputation model parameters is propagated through to the resulting imputations (26)). For example, Stata command *mi impute chained* (version 17 (51)) and R package *mice* (version 3.14.0) specify a uniform prior (e.g., prior *p(v, ς) ∝* ^*1*^*/* _*ς*_ for a linear regression with coefficients *v* and variance parameter *ς*, and prior *p (v) ∝* 1 for a logistic regression with coefficients *v*) (52,53).

To implement a probabilistic bias analysis using NARFCS we must specify a prior distribution for *δ*^*NARFCS*^. In keeping with Bayesian SM, we use prior *p(δ*^*NARFCS*^ *)∼N (μ*^*NARFCS*^, *σ*^*NARFCS*^ ) with *μ*^*NARFCS*^ and *σ*^*NARFCS*^ set to values based on external information.

#### Monte Carlo implementation

The Monte Carlo implementation of a probabilistic bias analysis repeatedly samples directly from the prior distribution for *δ*^*NARFCS*^ before fitting the bias model. Therefore, no sampled values of *δ*^*NARFCS*^ are rejected due to incompatibility with the observed data. Using the NARFCS bias model, we generate a Monte Carlo frequency distribution of bias-adjusted estimates of *β*_*X*_ by repeatedly carrying out the following steps *S(S>*1) times: for *S=* 1, *…, S*

i. Randomly draw a value for the bias parameter directly from its prior distribution, *δ*^*NARFCS(s)*^*∼ N(μ*^*NARFCS*^, *σ*^*NARFCS*^).
ii. Multiply impute the observed data *K(K ≥*1) times using the NARFCS bias model with the bias parameter fixed at *δ*^*NARFCS*(*s*)^. Fit the substantive analysis separately to each imputed dataset using maximum likelihood estimation and combine the multiple sets of results for *β*_*X*_ using Rubin’s rules (54). Let 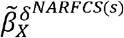 and 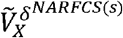 denote the combined estimate of *β*_*X*_ and accompanying variance, respectively.
iii. Randomly sample 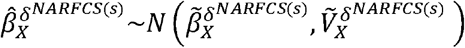 to incorporate random sampling error.

After *S* steps, we compute the median, 2.5^th^ and 97.5^th^ percentiles of the frequency distribution of 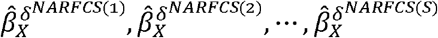 to obtain our Monte Carlo NARFCS bias-adjusted point and interval estimates of *β*_*X*_ . Monte Carlo NARFCS was implemented in R using the NARFCS extension to *mice* (55) and in Stata using the ‘offset’ option within the *mi impute* command.

### Simulation study design

We compared the performance of Monte Carlo NARFCS with that of Bayesian SM in an extreme situation of a large proportion of missing data under a very strong MNAR mechanism. We evaluated these methods when the prior distribution for the bias parameter was (i) inaccurate and imprecise, (ii) accurate and reasonably precise, and (iii) accurate and very precise. We repeated the simulation study for (1) *β*_*X*_ *=*0 and *β*_*X*_ *=* ln 3 and (2) two data generating models: based on the selection model framework (SM data generating model) and the pattern-mixture model framework (PMM data generating model). For all combinations of the simulation settings, we generated 1000 simulated data sets, each with 100,000 observations for the full sample.

#### Generation of the complete data

The simulation study was based on the hypothetical example described above with the exception that *Z= Z*_1_, *Z*_2_, *Z*_3_) denotes three fully observed confounders and *A= (A*_1_, *A*_2_) denotes two fully observed auxiliary variables. Exposure *X* and *Z*_2_ were continuous variables with mean 0 and standard deviation of 1, and the remaining variables were binary (*Z*_1_, *Z*_3_, *A*, outcome *Y*, partially observed confounder *W*, partially observed auxiliary *D* and missingness indicators *M*^*Y*^,*M*^*X*^, *M*^*W*^, and *M*^*D*^).

First, we simulated (complete) data on *X, Y, Z, W, A, D*, and *M*^*Y*^ from their joint distribution factorised into a series univariate regressions: logistic regression for *Y, Z*_1_, *Z*_3_, *W, A, D*, and *M*^*Y*^, and linear regression for *X* and *Z*_3_. We considered two factorisations of this joint distribution, with the factorisation for the SM and PMM data generating models chosen to resemble the bias model of Bayesian SM and Monte Carlo NARFCS, respectively. The equations for the SM and PMM data generating models, and their parameter values, are reported in sections 1.1 and 1.2 of the supplementary materials.

Most of the parameter values of the SM data generating model were set to estimates from an analysis of a real dataset (our motivating example, described in the next section). The marginal prevalence of *Y* was fixed at 5% for both the *β*_*X*_ *=*0 and *β*_*X*_ *=* ln 3 scenarios. The equivalent parameter values of the PMM data generating model were unknown. Following White et al (56), we estimated these parameter values by fitting the PMM data generating model to a dataset of 50,000,000 observations simulated under the SM data generating model.

#### Generation of the missing data

Following generation of the complete data, which included missingness indicator *M*^*Y*^, values of *Y* were set to missing when *M*^*Y*^ *=* 1. Missing data for *X, W*, and *D* were subsequently generated independently of each other and of *Y* using the following missingness mechanisms of MAR given fully observed variables:

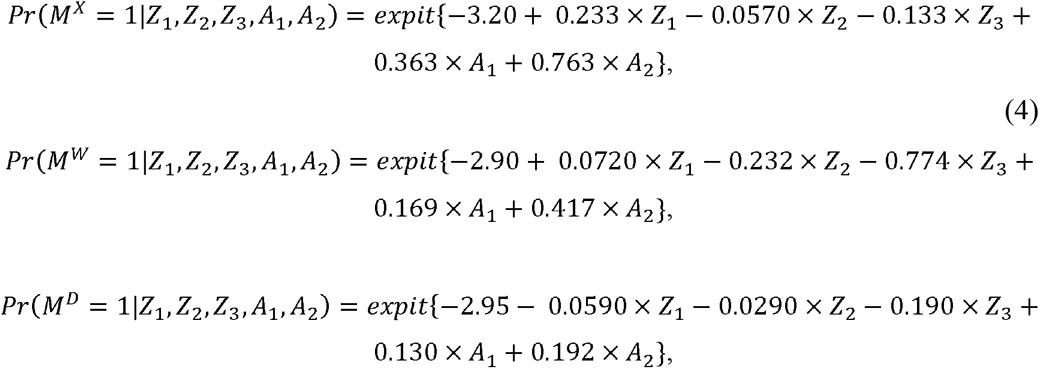

where all parameter values were derived from the observed data of our motivating example. These missingness mechanisms were the same for both the *β*_*X*_ *=*0 and *β*_*X*_ *=* ln 3 scenarios and the SM and PMM data generating models, resulting in a non-monotone missingness pattern. Close to 5% of the observations of *X, W*, and *D* were set as missing (e.g., *Pr(M*^*X*^ = 1)∼5%).

#### Missing data methods and evaluation

Probabilistic bias analyses, Bayesian SM and Monte Carlo NARFCS, were implemented as described previously (supplementary sections 1.3 and 1.4 show the extended versions of their bias models replacing *Z* with *Z*_1_, *Z*_2_, *Z*_3_ and *A* with *A*_1_, *A*_2_). Bayesian SM was applied with 50,000 iterations, of which 5,000 were burn-in iterations. This decision was based on running standard convergence checks (42) on one randomly selected dataset. Monte Carlo NARFCS was applied with 10,000 Monte Carlo steps and single imputation within each step. To assess sensitivity to the number of Monte Carlo steps and imputed datasets, we also conducted Monte Carlo NARFCS using 10,000 Monte Carlo steps with five imputations, and 5,000 Monte Carlo steps with single imputation. The number of burn-in iterations of the FCS algorithm was always set to 10. We applied Bayesian SM and Monte Carlo NARFCS with three different priors for the bias parameter: (i) vague prior *N(*0,100), (ii) informative prior *N(truth*, 4), and (iii) very informative prior *N(truth*, 1), where *truth* denotes the true value of the bias parameter. Note that the true value of *δ*^*NARFCS*^ was unknown (since it was not a parameter of either data generating model) and so we instead used an estimate of *δ*^*NARFCS*^ obtained by fitting the relevant conditional regression of the NARFCS bias model (3) to a simulated dataset of size 50,000,000 observations.

We compared Bayesian SM and Monte Carlo NARFCS to a CCA, MI and IPW. We applied MI using FCS imputation with 10 burn-in iterations and 50 imputations, imputing the binary and continuous variables using logistic and linear regressions, respectively (see supplementary section 1.5 for further details). We applied IPW with unstabilised and stabilised weights and considered three different weighting models (see supplementary section 1.6 for further details). Note that the weights could not be estimated for participants with missing data on the predictors of the weighting models and so these observations (which represented <0.5% of the full sample) were excluded from the IPW analysis. Standard errors for IPW were conservatively estimated using a sandwich estimator, treating the weights as known (30).

The estimand of interest was the exposure effect *β*_*X*_. For the SM data generating model, the true value of *β*_*X*_ was known as it was a parameter of this model, whilst for the PMM data generating model, a value for *β*_*X*_ was computed by fitting the substantive analysis to a dataset of size 50,000,000 before data deletion. Performance measures of interest were bias, empirical, and model-based standard errors, and 95% confidence interval (CI) coverage of estimates of *β*_*X*_. We used Stata version 17.0 (51) to generate the data. The remaining methods were conducted in R 4.1.0 (57). Bayesian SM and Monte Carlo NARFCS were applied using high performance computing for parallel processing (58) across the simulated datasets. R package *rsimsum* (version 0.11.3) (59) was used to compute the simulation results.

### Motivating example

The motivating example for our simulation study is a previously described study where the substantive analysis of interest is a logistic regression of body mass index (BMI) on SARS-CoV-2 infection (0 not infected, 1 infected) adjusted for confounders age, sex (0 female, 1 male), university degree (0 no, 1 yes), and current smoker (0 no, 1 yes) (60). This motivating example illustrates derivation of an informative prior for *δ*^*SM*^ and *δ*^*NARFCS*^ and the challenges of applying the fully Bayesian approach to a dataset with a large amount of missing data. As this is an illustrative example, we have ignored other potential sources of bias (such as selection bias due to non-random participation in UK Biobank (61)), and we have only considered a small number of confounders of the outcome–exposure relationship.

#### Motivating case study

Using data from the UK Biobank study (UKB) (61), we define our target population as middle aged and elderly adults (aged 47 – 86, with close to 75% of participants aged 61 or older) resident and alive in England on 1^st^ January 2020. Active SARS-CoV-2 infection was defined as either a positive SARS-CoV-2 PCR test (from linked Public Health England data) or COVID-19 recorded on a death certificate between 1^st^ January 2020 and 18^th^ May 2020 (i.e., the date mass testing became available in the UK; (62)). Testing for SARS-CoV-2 was highly restricted during this period and so data on SARS-CoV-2 infection were missing for the majority of participants (over 98% of the study sample). Data on SARS-CoV-2 infection were suspected to be MNAR since testing among the majority of the UK population (i.e., non-healthcare workers) was mainly restricted to those who experienced symptoms of COVID-19 (63). Observed factors associated with the chance of being tested in UKB included having higher BMI, being a current smoker, having a pre-existing condition (such as asthma, diabetes, or hypertension), being female, and having a university degree or higher (60).

Among the 445,377 participants included in the UKB study, we excluded 24,465 (5.49%) participants who died before 2020 and 65 (0.0146%) who were not tested for SARS-CoV-2 but were diagnosed with COVID-19 post-mortem. Of the remaining 420,847 participants eligible for analysis, 405,174 (96.3%) were missing the outcome only, 10,870 (2.58%) were missing the outcome and at least one covariate (BMI, smoker, or degree), 4,610 (1.10%) had complete data and 193 (0.0459%) had an observed outcome but were missing at least one covariate (supplementary table 3). Confounders age and sex, and auxiliary variables asthma, diabetes, and hypertension were fully observed. Figure 2 shows the m-DAG for this motivating example based on subject-matter knowledge and our investigations of observed predictors of missingness (supplementary tables 4 and 5). In the absence of contrary information, we allow the covariate data to be MNAR depending on its own missing values. Although outcome, SARS-CoV-2, cannot be a direct cause of missingness of the covariate data, since the outcome occurred many years after the covariates were recorded, there may be common causes of SARS-CoV-2 and the missing covariate data. We assume there were no unmeasured common causes after accounting for age, sex, degree, smoker, BMI, asthma, diabetes, and hypertension.

**Figure 2:**
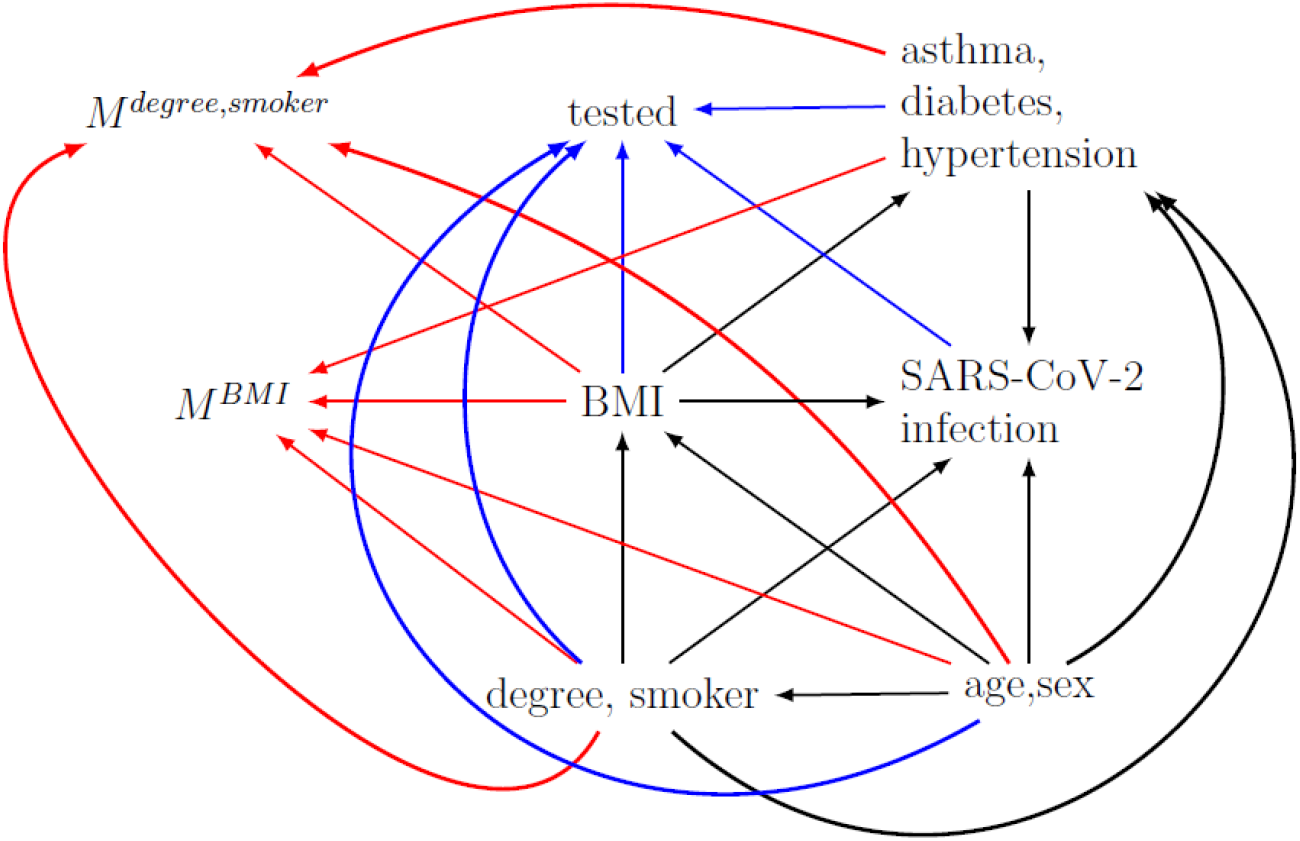
Missingness directed acyclic graph for the UK Biobank example. Black edges depict the assumed relationships in the fully observed data between the outcome (SARS-CoV-2 infection), exposure (body mass index (BMI)), confounders (age, sex, degree, and smoker), and auxiliary variables (asthma, diabetes, and hypertension). Tested, M^BMI^, and M^degree,smoker^ denote missingness indicators for the outcome, exposure, and confounders, respectively. Blue and red edges depict the missingness mechanisms of the outcome and covariates (exposure and confounders), respectively. Note, we have not included all edges between the variables.

#### Statistical analyses

We analysed the data using CCA, MI, IPW, Bayesian SM, Monte Carlo NARFCS, and a “population-based comparison group approach” where untested participants were assumed to be not infected with SARS-CoV-2 (64,65,66). See section 2.2 of the supplementary materials for details of the imputation, weighting, and bias models fitted. Due to convergence problems encountered when applying Bayesian SM to the full data, we restricted all analyses to the 409,784 participants with complete data on the covariates. This simplified the imputation, weighting, and bias models (e.g., removal of the regression models for BMI, degree, and smoker from the Bayesian SM bias model), thus reducing the number of parameters to be estimated. Given the small percentage of dropped participants (the majority of which had a missing outcome), the characteristics of the full sample and the restricted sample were virtually the same (supplementary table 6). In keeping with the preceding paper (60), and to improve the efficiency of MCMC sampling by reducing autocorrelation in the chains, each continuous variable (age and BMI) was standardized by subtracting its observed mean and dividing by its observed standard deviation. These standardised variables were used in all analyses. We applied MI with 50 imputed datasets, Bayesian SM using 50,000 MCMC iterations (including 5,000 burn-in iterations), and Monte Carlo NARFCS with 10,000 Monte Carlo steps and single imputation. Bayesian SM and Monte Carlo NARFCS were applied using an informative prior for *δ*^*SM*^ and *δ*^*NARFCS*^, respectively.

#### Derivation of the informative priors for δ^SM^ and δ^NARFCS^

The hyperparameters of the informative priors *p(δ*^*SM*^*)∼N(μ*^*SM*^, *σ*^*SM*^) and *p(δ*^*NARFCS*^ *)∼N(μ*^*NARFCS*^, *σ*^*NARFCS*^ ) were derived from published results of the REal-time Assessment of Community Transmission-2 (REACT-2) national study (67). The REACT-2 study sent home-based SARS-CoV-2 antibody test kits to over 100,000 randomly sampled adults living in England between 20^th^ June and 13^th^ July 2020. Among 65–74-year-olds (similar age range to our study), SARS-CoV-2 antibody prevalence was estimated to be 3.2% [95% CI 2.8–3.6%] (67) by mid-July 2020.

Bias parameters *δ*^*SM*^ and *δ*^*NARFCS*^ are conditional parameters on the log-odds scale. So, we used an algorithm from Tompsett et al (27) to compute approximate values of *δ*^*SM*^ and *δ*^*NARFCS*^ calibrated to marginal prevalences of SARS-CoV-2 infection. For prior *p(δ*^*NARFCS*^ *)∼N(μ*^*NARFCS*^, *σ*^*NARFCS*^ ), we set *μ*^*NARFCS*^ *= −*2.6 (the value of *δ*^*NARFCS*^ calibrated to a prevalence of 3.2%) and set *σ*^*NARFCS*^ *=* 0.22^2^ such that 95% of the sampled values of *δ*^*NARFCS*^ were expected to be between -3.0 and -2.2 (which were the values of *δ*^*NARFCS*^ calibrated to prevalences of approximately 2.2% and 4.2%, respectively). Note that we allowed for additional uncertainty because the prevalence of infection was unknown in our UKB study. The comparable prior for Bayesian SM was *p(δ*^*SM*^*)∼N*(2.6, 0.22^2^). Further details, including the algorithm used, are described in section 2.2.1 of the supplementary materials.

## Results

### Simulation study results

When there were no missing data, the full data estimate of *β*_*X*_ was unbiased and CI coverage was close to the nominal level in all scenarios. Figure 3 shows the bias and coverage of estimating *β*_*X*_ in the presence of missing data using different missing data methods when the true value of *β*_*X*_ was ln 3 and 0 and the data were generated using the SM data generating model (detailed results reported in supplementary tables 8 and 9). There was substantial bias and severe CI under-coverage for the CCA estimates, with similar levels of bias for *β*_*X*_ *=* ln 3 and *β*_*X*_ *=*0 but slightly higher CI coverage for *β*_*X*_ *=* ln 3 due to larger (empirical and model-based) standard errors. Note that in both scenarios, the CCA CIs were “centred” around a biased point estimate whilst the model-based standard errors were unbiased. Given the same level of bias in the point estimates, the wider CIs of the *β*_*X*_ *=* ln 3 scenario were more likely to contain the true value of *β*_*X*_ than the narrower confidence intervals of the *β*_*X*_ *=*0 scenario. When *β*_*X*_ *=* 0, MI and IPW had broadly comparable levels of bias and CI coverage to CCA. However, when *β*_*X*_ = ln 3, the bias of the MI estimates was noticeably larger than that of CCA. This was likely due to amplification of the bias (resulting from incorrect assumptions about the missingness mechanism) caused by including variables in the imputation model that were strongly predictive of *Y* (33) (see section 3.2 for further details). Also, when *β*_*X*_ = ln 3, despite similar levels of bias the CI coverage of IPW with unstabilised weights was substantially larger than that of CCA. The inflated standard errors of IPW were due to the presence of large weights, which were driven by the strong relationships between *Y* and *A*_1_, *A*_2_, and *D*. Hence the problem of inflated standard errors only occurred for the *β*_*X*_ *=* ln 3 scenario since these relationships were null for the *β*_*X*_ *=*0 scenario. Using IPW with stabilised weights reduced the problem of inflated empirical standard errors but resulted in a higher level of bias because lower variability and skewness in the estimates of *β*_*X*_ led to fewer over-estimates compensating for the negative bias induced by the missing data. Similar results were obtained for the other weighting models (supplementary table 10) and in the simpler setting of missing *Y* only (see supplementary section 3.3 for further details).

**Figure 3:**
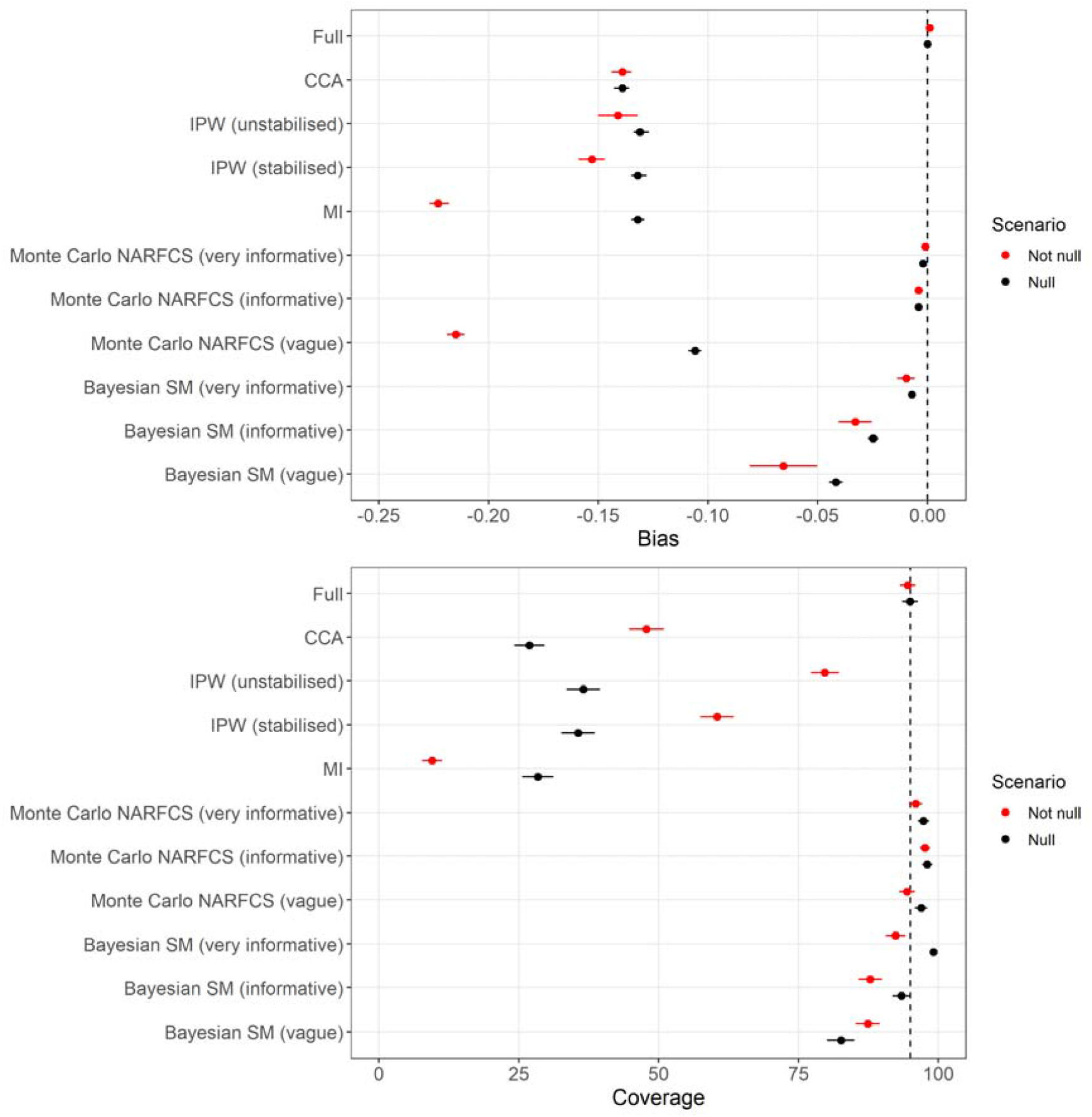
Bias and 95% confidence interval coverage of exposure effect, *β*_*X*_, according to the not null (*β*_*X*_ *=* ln 3) and null (*β*_*X*_ *=* 0) scenarios for data generated using SM data generating model . Error bars denote 95% Monte Carlo intervals, and the vertical dashed line denotes zero bias (top) and nominal coverage (bottom). Results for Bayesian SM were based on 926-928 simulated datasets; the remaining methods were based on 1,000 simulated datasets.

For both the *β*_*X*_ *=* ln 3 and *β*_*X*_ =0 scenarios, there was negligible bias when applying Monte Carlo NARFCS with an informative or very informative prior. (See below for our comments on the CI overcoverage). Applying Monte Carlo NARFCS with a vague prior resulted in biased estimates where the level of bias was slightly lower than that of CCA for the *β*_*X*_ = ln 0 scenario but higher for the *β*_*X*_ *=* ln 3 scenario (and comparable to that of MI). In accordance with MI, the higher level of bias for the *β*_*X*_ = ln 3 scenario was likely due to the auxiliary variables amplifying the bias from misspecification of the missingness mechanism (see supplementary section 3.2 for further details). Despite the (relatively) high level of bias, CI coverage was nominal due to the imprecision of the vague prior. Very similar results were obtained when applying Monte Carlo NARFCS with 10,000 Monte Carlo steps with 5 imputations and 5,000 Monte Carlo steps with single imputation (supplementary tables 15 and 16).

Method Bayesian SM failed to produce results for 72 to 74 simulated datasets (further details in supplementary section 3.5) whilst the other methods returned results for all 1,000 simulated datasets. Similar to Monte Carlo NARFCS, applying Bayesian SM with an informative or very informative prior resulted in minimal bias. However, compared to Monte Carlo NARFCS, Bayesian SM showed slightly higher levels of bias and inefficiency (i.e., larger empirical standard errors), leading to moderate levels of CI under-coverage. This seeming under-performance of Bayesian SM may have been due to the omitted estimates caused by nonconvergence in a small number of datasets (see section 3.5.2 of the supplementary materials). Unlike Monte Carlo NARFCS, Bayesian SM with a vague prior eliminated some of the bias in both the *β*_*X*_ *=* ln 3 and *β*_*X*_ *=*0 scenarios, with bias levels at least 50% lower than those of CCA. Also, the model-based standard errors of Bayesian SM were considerably smaller than those of Monte Carlo NARFCS. A likely explanation is that some information was gained from the Bayesian approach combining the prior for *δ*^*SM*^ with the observed data. Supporting this claim, we note that when applied with an *a priori* mean of 0 for *δ*^*SM*^, across the simulations the mean of the posterior estimates of *δ*^*SM*^ was 8.83 (95% Monte Carlo interval 8.31 to 8.96) and 6.36 (95% Monte Carlo interval 6.20 to 6.52) for the *β*_*X*_ *=* ln 3 and *β*_*X*_ *=*0 scenarios, respectively (where the true value was 7.85).

For both Bayesian SM and Monte Carlo NARFCS with (very) informative priors, there was CI overcoverage when the estimates of *β*_*X*_ were unbiased (or negligibly biased). This overcoverage was likely due to generating the data using a fixed value for the bias parameter (as opposed to sampling from an appropriate prior) which is known to lead to CI overcoverage when applying an analysis with an informative prior centred on the true value of the parameter (68); that is, were we to use a point-prior at the true value, we would expect to see nominal coverage (14). Also, this CI inflation will have led to higher-than-expected coverage levels for Bayesian SM in scenarios where bias was non-negligible.

Similar patterns were noted on the relative performances of the methods for data generated using the PMM data generating model (supplementary tables 18 and 19). For both data generating models and *β*_*X*_ *=* ln 3 and *β*_*X*_ *=*0 scenarios, Bayesian SM took substantially longer to run than Monte Carlo NARFCS with Monte Carlo NARFCS taking approximately 2 days per dataset in R (approximately 1 day per dataset in Stata) and Bayesian SM taking approximately 6 days per dataset.

### Results of the motivating example

Of the 409,784 participants included in our analysis with complete covariate data, 4,610 (1.12%) were tested for SARS-CoV-2, leaving 405,174 (98.9%) with a missing outcome. Out of the 4,610 participants tested for SARS-CoV-2, 1,317 (28.6%) tested positive. Figure 4 shows the results for the exposure odds ratio (i.e., odds ratio of SARS-CoV-2 infection per standard deviation increase in BMI) estimated using CCA, IPW, MI, Bayesian SM, Monte Carlo NARFCS, and the population-based comparison group approach. All analyses suggested that participants with a higher BMI tended to be at a higher risk of SARS-CoV-2 infection. The two probabilistic bias analyses, Bayesian SM and Monte Carlo NARFCS, gave similar results with slightly higher point estimates than CCA and the two MAR analyses, MI and IPW, although there was substantial overlap between the CIs of these methods. The results for the population-based comparison group approach were markedly different from those of the other methods.

**Figure 4:**
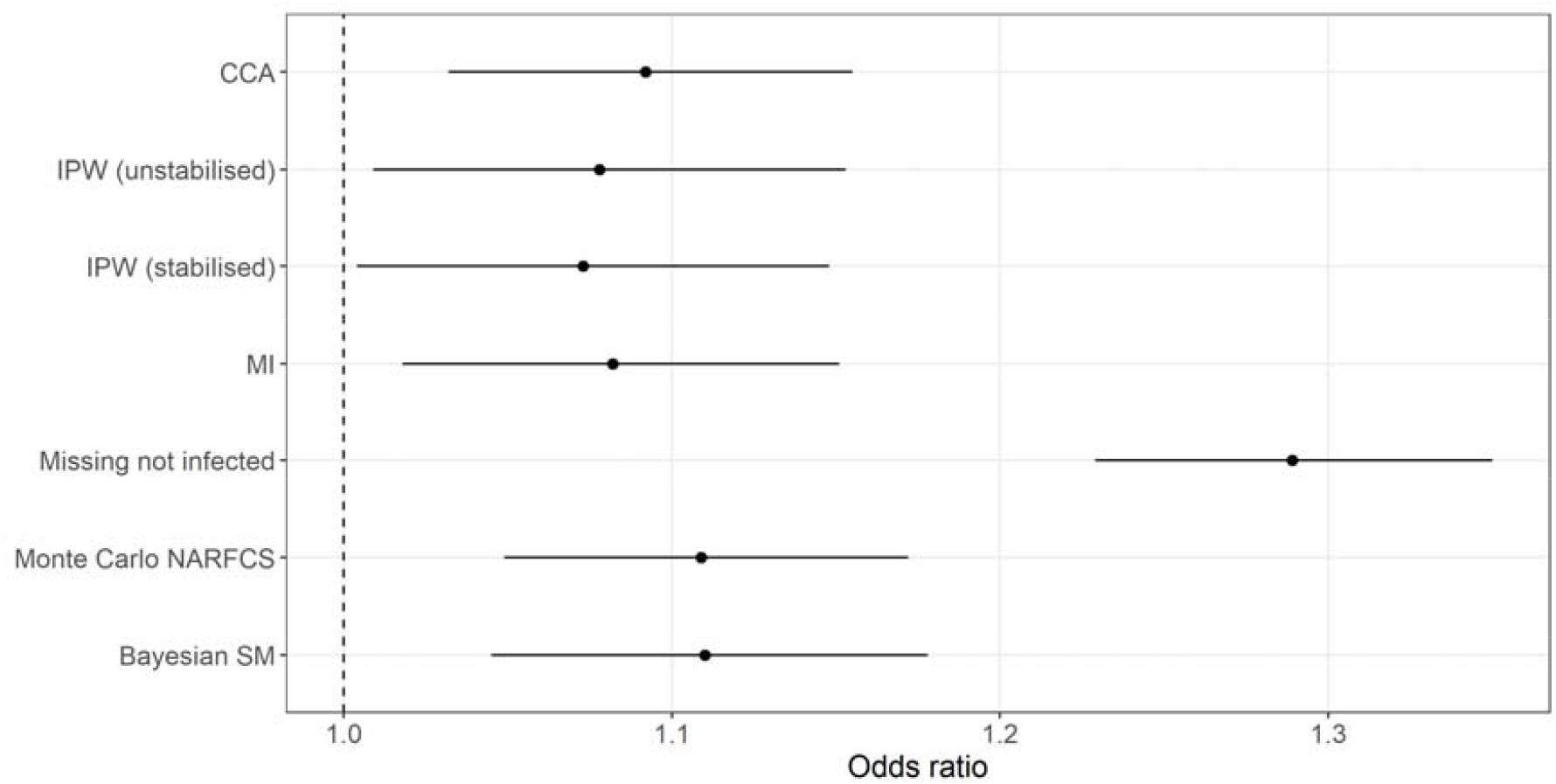
Forest plot of the results for exposure odds ratio, *exp{β*_*X*_*}*, estimated by complete case analysis (CCA), inverse probability weighting (with unstabilised and stabilised weights) and multiple imputation assuming missing at random (IPW and MI, respectively), population-based comparison group approach (Missing not infected), and the probabilistic bias analyses, Monte Carlo NARFCS and Bayesian SM. Dashed line denotes the null effect.

The patterns in the results were consistent with our prior knowledge that untested participants tended to have a lower BMI and were less likely to have experienced symptoms of SARSCoV-2 infection than tested participants. For example, under this missingness mechanism we expected that dropping untested participants (i.e., CCA) would lead to an underestimate of the exposure odds ratio (as demonstrated by the simulation study) and setting all untested participants as “not infected” would lead to an overestimate. The analyses based on the untested and tested participants (i.e., MI, Monte Carlo NARFCS, Bayesian SM, and the population-based comparison group approach) had similar levels of precision to the weighted and unweighted analyses of the tested participants (i.e., IPW and CCA). This was unsurprising given that (i) precision of binary outcome estimators is primarily determined by the number of cases (i.e., positive SARS-CoV-2 infections in our example) and (ii) for our study population and study period, the prevalence of SARS-CoV-2 infection was estimated to be relatively low (i.e., 3.2% [95% CI 2.8–3.6%] (67)) and so a large proportion of the untested participants were likely not infected with SARS-CoV-2. The distinct results of the population-based comparison group approach was due to the imposed extreme scenario which implied that the prevalence of infection in the study sample was only 0.32%.

## Discussion

We have illustrated the feasibility and practicality of conducting a probabilistic bias analysis to data MNAR when a large proportion of an outcome is missing under a strong MNAR mechanism. Our simulation study demonstrated that when reasonably accurate and precise information was provided about the bias parameter, the simpler, Monte Carlo NARFCS method performed as well as the more principled, fully Bayesian SM method. When very limited information was provided about the bias parameter, the fully Bayesian probabilistic bias analysis was able to eliminate most of the bias due to data MNAR while the Monte Carlo probabilistic bias analysis performed no better than the CCA and MAR implementations of IPW and MI. We have also shown how including auxiliary variables in an imputation model can amplify bias due to data MNAR even when their inclusion was expected to eliminate some of the bias due to missing data.

Monte Carlo NARFCS has two key strengths for non-specialist analysts: (1) the Monte Carlo implementation of a probabilistic bias analysis does not require knowledge about Bayesian inference and specialist statistical software, and (2) it calculates the bias-adjusted estimates using an MNAR-adaption of a popular and readily implemented imputation approach (FCS). However, both the Monte Carlo probabilistic bias analysis and FCS-type imputation have known theoretical weaknesses. Therefore, it is encouraging that Monte Carlo NARFCS can perform as well as the more principled Bayesian SM (i.e., with respect to the fully Bayesian analysis using a joint model for the multivariate distribution of the observed and missing data and the missingness mechanism). This is supported by previous research, which has established the robustness of FCS imputation to its theoretical weakness (that the joint distribution implied by the univariate regression models may not exist (49,50,69)). During the simulation study we experienced some minor technical difficulties with Bayesian SM.

However, these issues can be easily resolved when applying the method in practice. For example, nonconvergence would be identified using standard Bayesian diagnostic tools and resolved by running a longer burn-in, and failure of the Bayesian sampler could be rectified by using different starting values or switching to a different Monte Carlo algorithm. In keeping with McCandless and Gustafson (14), we found that applying a fully Bayesian probabilistic bias analysis using a vague prior for the bias parameter gained some information about the MNAR mechanism and consequently eliminated some of the bias due to missing data. This was likely due to the Bayesian process ruling out certain MNAR mechanisms (i.e., values of the bias parameter) incompatible with the observed data (14). In contrast, the Monte Carlo approach of sampling directly from the prior distribution of the bias parameter, irrespective of the observed data, meant that applying the Monte Carlo probabilistic bias analysis with a vague prior performed as badly as the MAR methods. Therefore, a fully Bayesian probabilistic bias analysis is recommended when there is limited information available about the bias parameters (i.e., MNAR mechanism in our case).

Another difference between the two probabilistic bias analyses is that the bias model of Bayesian SM is a selection model while that of Monte Carlo NARFCS is a pattern-mixture model. The advantage of the selection model framework (i.e., a model for the full data and a model for the missingness mechanism) is that it is coherent with our understanding of how the observed data arises and there is a logical separation of the parameters of interest from the bias parameters (11). However, others have argued that the bias parameters of the pattern-mixture model are usually easier to interpret and so this framework is more convenient for conducting bias analyses (12,70). In our applied example, the available external information was not ideally suited for the bias parameter of either the selection or pattern-mixture model and it was necessary to convert the external information about a marginal parameter to the required conditional parameter for both models. In general, the choice between the selection and pattern-mixture frameworks will depend on the aims of the analysis (e.g., is the missingness mechanism of primary interest) and the format of the available (external) information about the relationship between the variable and its chance of missingness.

Our simulation study has several limitations. First, our comparison of a Bayesian probabilistic bias analysis to a Monte Carlo probabilistic bias analysis also differed with respect to specification of the bias model. However, the primary objective of our study was to illustrate an easy to apply probabilistic bias analysis (for non-specialist analysts) and to compare it to a principled approach (with regards to its fully Bayesian implementation). Second, when simulating the data, we fixed the value of the bias parameter. Consequently, we would not expect nominal CI coverage for the Monte Carlo and Bayesian probabilistic bias analyses. However, we think it is reasonable to assume that the true bias parameter is fixed (as we do in our data generating models) and we allow for the expected extra uncertainty (i.e., over-coverage). Also, we think it is an advantage for a probabilistic bias analysis approach to incorporate extra uncertainty since the true value of the bias parameter will always be unknown. We focus on what Rubin terms confidence validity (i.e., intervals that cover at least nominally) rather than randomisation validity (i.e., intervals that cover exactly nominally) (71). Related to this, our approach exhibited coverage of 95% even when the prior for the bias parameter was not centred on the fixed true value.

Third, we only explored a small number of scenarios because of the time it took to run each probabilistic bias analysis in a large data setting (typical of cohort studies). To achieve our objective of evaluating the robustness of Monte Carlo NARFCS using a small-scale simulation study, we considered an extreme missingness setting (i.e., large proportion of missing data with a strong MNAR mechanism).

In summary, when information is available on the bias parameter, our simpler Monte Carlo probabilistic bias analysis is a viable alternative to a fully Bayesian probabilistic bias analysis. However, when limited information is available, a fully Bayesian probabilistic bias analysis is recommended over the Monte Carlo approach. Both proposed probabilistic bias analyses can be implemented in standard statistical software and can be extended to deal with multiple MNAR mechanisms. By illustrating two different types of probabilistic bias analyses and providing code to replicate them, we hope to encourage the increased adoption of such bias analyses in epidemiological research. Finally, in keeping with (33,34), we caution careful consideration of the choice of auxiliary variables when applying MI where data may be MNAR.

## Data Availability

The software code to generate the simulated datasets analysed during the simulated study are available in the COVIDITY_ProbQBA repository, https://github.com/MRCIEU/COVIDITY_ProbQBA. The UK Biobank study dataset analysed during the current study is available from the UK Biobank Access Management Team (https://www.ukbiobank.ac.uk/learn-more-about-uk-biobank/contact-us) but restrictions may apply to the availability of these data, which were used under license for the current study, and so are not publicly available. All methods discussed in this paper can be implemented using the provided software code available from the COVIDITY_ProbQBA repository, https://github.com/MRCIEU/COVIDITY_ProbQBA.

## Abbreviations

BMI: body mass index
CCA: complete case analysis
FCS: fully conditional specification
IPW: inverse probability weighting
MAR: missing at random
MCMC: Markov Chain Monte Carlo
MI: multiple imputation
MNAR: missing not at random
NARFCS: not-at-random fully conditional specification
PMM: pattern mixture model
REACT-2: REal-time Assessment of Community Transmission-2
SM: selection model
UKB: UK Biobank study

## Acknowledgements

Not applicable.

## Funding

This work was supported by the Bristol British Heart Foundation (BHF) Accelerator Award (AA/18/ 7/34219), the University of Bristol and Medical Research Council (MRC) Integrative Epidemiology Unit (MC_UU_00032/01, 02 & 05), the BHF-National Institute of Health Research (NIHR) COVIDITY flagship project, the John Templeton Foundation (61917), and the Wellcome Trust and Royal Society (215408/Z/19/Z). TPM was funded by the UKRI Medical Research Council, grant number MC_UU_00004/09. The computation work was carried out using the computational facilities of the Advanced Computing Research Centre, University of Bristol - http://www.bristol.ac.uk/acrc/.

## Authors’ contributions

Authors EK, DM-S, GLC, CYS, TPM, ARC, AF-S, MCB, GJG, LACM and RAH designed the study with critical review from KT, DL AND GDS. EK, DM-S and GLC performed the simulation study and statistical analyses under the supervision of RAH, CYS and TPM. EK, DM-S, GLC and RAH drafted the paper with input from the remaining authors. All authors were responsible for critical revision of the manuscript and have approved the final version to be published.

## Ethics approval and consent to participate

For the simulation study, data were completely simulated, which did not require approval from an ethics committee or consent from participants. UKB received ethical approval from the UK National Health Service’s National Research Ethics Service (ref. 11/NW/0382). All participants provided written and informed consent for data collection, analysis, and record linkage. This research was conducted under UKB application number 16729.

## Competing interests

TPM has received consultancy fees from: Bayer Healthcare Pharmaceuticals, Alliance Pharmaceuticals, Gilead Sciences, and Kite Pharmaceuticals. Since January 2023, ARC has been an employee of Novo Nordisk Research Centre Oxford, which is not related to the current work and had no involvement in the decision to publish. The remaining authors declare that they have no competing interests.

## Consent for publication

Not applicable.

## Notes

### Author Declarations

UK Biobank received ethical approval from the UK National Health Service's National Research Ethics Service (ref. 11/NW/0382). All participants provided written and informed consent for data collection, analysis, and record linkage. This research was conducted under UKB application number 16729.

