## Supplementary materials for "Accounting for bias due to outcome data missing not at random: comparison and illustration of two approaches to probabilistic bias analysis: a simulation study"

### **1 Additional details of the simulation study design**

#### **1.1 Selection model data generating model**

The selection model (SM ) data generating model was defined as follows:

$Z_{1} \sim Bernoulli\left( 0.457 \right)$, $Z_{2}\sim N\left( -0.021+0.046\times Z_{1},1^{2} \right)$, $Pr\left( Z_{3}=1 \right)=expit\left\{ -0.795+ 0.140\times Z_{1}-0.272\times Z_{2} \right\}$,

$Pr\left( W=1 \right)=expit\left\{ -2.221+ 0.406\times Z_{1}-0.303\times Z_{2}-0.600\times Z_{3} \right\}$,

$X\sim N\left( 0.018+0.168\times Z_{1}+0.024\times Z_{2}-0.260\times Z_{3}-0.123\times W,{0.987}^{2} \right)$,

$Pr\left( Y=1 \right)=expit\left\{ \beta_{0}+ 0.311\times Z_{1}+0.043\times Z_{2}-0.318\times Z_{3}+0.213\times W+\beta_{X}\times X \right\}$,

$Pr\left( A_{1}=1 \right)=expit\left\{ -1.186+ 0.069\times Z_{1}+0.131\times Z_{2}-0.091\times Z_{3}+0.413\times W+0.188\times X+\theta_{Y}\times Y \right\}$,

$Pr\left( A_{2}=1 \right)=expit\left\{ -3.097+ 0.617\times Z_{1}+0.426\times Z_{2}-0.257\times Z_{3}+0.331\times W+0.756\times X+o_{Y}\times Y+0.497\times A_{1} \right\}$,

$Pr\left( D=1 \right)=expit\left\{ -0.895+ 0.349\times Z_{1}+0.649\times Z_{2}-0.200\times Z_{3}+0.094\times W+0.527\times X+\omega_{Y}\times Y+0.380\times A_{1}+1.374\times A_{2} \right\}$,

$Pr\left( M^{Y}=1 \right)=expit\left\{ \psi_{0}- 0.013\times X-7.849\times Y-0.991\times A_{1}-0.519\times A_{2}-0.441\times D \right\}$,

where $expit\left\{ k \right\}=\frac{exp\left\{ k \right\}}{1+exp\left\{ k \right\}}$ and the bias parameter $\delta^{SM}=-7.849;$that is, the coefficient of $Y$ in logistic regression $Pr\left( M^{Y}=1|X,Y,A_{1},A_{2},D \right)$. The null ($\beta_{X}=0$) and not null ($\beta_{X}=\ln3$) exposure effect scenarios were generated by setting parameters $\beta_{0},\beta_{X},\theta_{Y},o_{Y}$*,* $\omega_{Y},$ and $\psi_{0}$ to the values reported in supplementary table 1 (intercept terms chosen to maintain P$r\left( Y=1 \right)$ and P$r\left( M^{Y}=1 \right)$ at 5%). The remaining parameters, including bias parameter $\delta^{SM}$, were fixed to the values shown above. Note that in the null scenario, $Y$ was independent of $A_{1},A_{2}$ and $D$ (given $Z_{1},Z_{2},Z_{3},W,X$); hence $\theta_{Y},o_{Y}$ and $\omega_{Y}$ were set to 0.

**Supplementary table 1:** Parameter values of the selection data generating model according to the null and not null exposure effect scenarios

| **Scenario** | $\boldsymbol{\beta}_{\boldsymbol{0}}$ | $\boldsymbol{\beta}_{\boldsymbol{X}}$ | $\boldsymbol{\theta}_{\boldsymbol{Y}}$ | $\boldsymbol{o}_{\boldsymbol{Y}}$ | $\boldsymbol{\omega}_{\boldsymbol{Y}}$ | $\boldsymbol{\psi}_{\boldsymbol{0}}$ |
| --- | --- | --- | --- | --- | --- | --- |
| Not null exposure effect | -3.5 | $\ln3$ | 1.617 | 1.228 | 1.011 | 6.3 |
| Null exposure effect | -3 | 0 | 0 | 0 | 0 | 6 |

#### **1.2 Pattern-mixture model data generating model**

The pattern-mixture model (PMM) data generating model was defined as follows:

$Z_{1} \sim Bernoulli\left( 0.457 \right)$, $Z_{2} \sim N\left( -0.021+0.046\times Z_{1},1^{2} \right)$, $Pr\left( Z_{3}=1 \right)=expit\left\{ -0.795+ 0.140\times Z_{1}-0.272\times Z_{2} \right\}$,

$Pr\left( W=1 \right)=expit\left\{ -2.221+ 0.406\times Z_{1}-0.303\times Z_{2}-0.600\times Z_{3} \right\}$,

$X\sim N\left( 0.018+0.168\times Z_{1}+0.024\times Z_{2}-0.260\times Z_{3}-0.123\times W,{0.987}^{2} \right)$,

$Pr\left( M^{Y}=1 \right)= expit\left\{ \psi_{0}^{PMM}+\psi_{Z_{1}}^{PMM}Z_{1}+\psi_{Z_{2}}^{PMM}Z_{2}+\psi_{Z_{3}}^{PMM}Z_{3}+\psi_{W}^{PMM}W+\psi_{X}^{PMM}X \right\}$,

$Pr\left( Y=1 \right)$ $= expit\left\{ \kappa_{0}^{PMM}+\kappa_{Z_{1}}^{PMM}Z_{1}+\kappa_{Z_{2}}^{PMM}Z_{2}+\kappa_{Z_{3}}^{PMM}Z_{3}+\kappa_{W}^{PMM}W+\kappa_{X}^{PMM}X +\kappa_{M^{Y}}^{PMM}M^{Y} \right\}$,

$Pr\left( A_{1}=1 \right)=expit\left\{ \theta_{0}^{PMM}+\theta_{Z_{1}}^{PMM}Z_{1}+\theta_{Z_{2}}^{PMM}Z_{2}+\theta_{Z_{3}}^{PMM}Z_{3}+\theta_{W}^{PMM}W+\theta_{X}^{PMM}X+\theta_{Y}^{PMM}Y+\theta_{M^{Y}}^{PMM}M^{Y} \right\}$,

$Pr\left( A_{2}=1 \right)=expit\left\{ \alpha_{0}^{PMM}+\alpha_{Z_{1}}^{PMM}Z_{1}+\alpha_{Z_{2}}^{PMM}Z_{2}+\alpha_{Z_{3}}^{PMM}Z_{3}+\alpha_{W}^{PMM}W+\alpha_{X}^{PMM}X+\alpha_{Y}^{PMM}Y+\alpha_{M^{Y}}^{PMM}M^{Y}+\alpha_{A_{1}}^{PMM}A_{1} \right\}$,

$Pr\left( D=1 \right)=expit\left\{ \omega_{0}^{PMM}+\omega_{Z_{1}}^{PMM}Z_{1}+\omega_{Z_{2}}^{PMM}Z_{2}+\omega_{Z_{3}}^{PMM}Z_{3}+\omega_{W}^{PMM}W+\omega_{X}^{PMM}X+\omega_{Y}^{PMM}Y+\omega_{M^{Y}}^{PMM}M^{Y}+\omega_{A_{1}}^{PMM}A_{1}+\omega_{A_{2}}^{PMM}A_{2} \right\}$,

where $expit\left\{ k \right\}=\frac{exp\left\{ k \right\}}{1+exp\left\{ k \right\}}$. Variables $Z_{1},Z_{2},Z_{3},$ and $X$ were generated exactly as described for the SM data generating model. For the remaining variables, parameters of the data generating regression models were determined by fitting the models to a dataset of 50,000,000 observations simulated under the SM data generating model. Note that, bias parameter $\delta^{NARFCS}$ is the coefficient of $M^{Y}$ from logistic regression $Y$ given $Z_{1},Z_{2},Z_{3},W,X,A_{1},A_{2},D,M^{Y},M^{X},M^{W},M^{D}$ and so is not equal to coefficient $\kappa_{M^{Y}}^{PMM}$ from logistic regression $Y$ given $Z_{1},Z_{2},Z_{3},W,X,A_{1},A_{2},D,M^{Y}$.

#### **1.3 Bias model of Bayesian SM**

The bias model of the Bayesian SM approach fitted to the simulated data consists of the following regression models:

$$Pr\left( W=1 \right)\sim expit\left\{ \eta_{0}+\eta_{Z_{1}}Z_{1}+\eta_{Z_{2}}Z_{2}+\eta_{Z_{3}}Z_{3} \right\}, X\sim N\left( \zeta_{0}+\zeta_{W}W+\zeta_{Z_{1}}Z_{1}+\zeta_{Z_{2}}Z_{2}+\zeta_{Z_{3}}Z_{3},\xi^{2} \right),$$

$Pr\left( Y=1 \right)\sim expit\left\{ \beta_{0}+\beta_{X}X+\beta_{W}W+\beta_{Z_{1}}Z_{1}+\beta_{Z_{2}}Z_{2}+\beta_{Z_{3}}Z_{3} \right\},$

$$Pr\left( A_{1}=1 \right)\sim expit\left\{ \theta_{0}+\theta_{Y}Y+\theta_{X}X+\theta_{W}W+\theta_{Z_{1}}Z_{1}+\theta_{Z_{2}}Z_{2}+\theta_{Z_{3}}Z_{3} \right\},$$

$$Pr\left( A_{2}=1 \right)\sim expit\left\{ o_{0}+o_{A_{1}}A_{1}+o_{Y}Y+o_{X}X+o_{W}W+o_{Z_{1}}Z_{1}+o_{Z_{2}}Z_{2}+o_{Z_{3}}Z_{3} \right\},$$

$$Pr\left( D=1 \right)\sim expit\left\{ \omega_{0}+\omega_{A_{1}}A_{1}+\omega_{A_{2}}A_{2}+\omega_{Y}Y+\omega_{X}X+\omega_{W}W+\omega_{Z_{1}}Z_{1}+\omega_{Z_{2}}Z_{2}+\omega_{Z_{3}}Z_{3} \right\},$$

$$Pr\left( M^{Y}=1 \right)\sim expit\left\{ \psi_{0}+\psi_{D}D+\psi_{A_{1}}A_{1}+\psi_{A_{2}}A_{2}+\delta^{SM}Y+\psi_{X}X \right\}.$$

#### **1.4 Bias model of Monte Carlo NARFCS**

The bias model of the Monte Carlo NARFCS approach fitted to the simulated data consists of the following regression models:

$$Pr\left( W=1 \right)\sim expit\left\{ \alpha_{0}+\alpha_{Y}Y+\alpha_{X}X+\alpha_{D}D+\alpha_{A_{1}}A_{1}+\alpha_{A_{2}}A_{2}+\alpha_{Z_{1}}Z_{1}+\alpha_{Z_{2}}Z_{2}+\alpha_{Z_{3}}Z_{3}+\alpha_{M^{Y}}M^{Y}+\alpha_{M^{X}}M^{X}+\alpha_{M^{D}}M^{D} \right\},$$

$$X\sim N\left( \gamma_{0}+\gamma_{Y}Y+\gamma_{W}W+\gamma_{D}D+\gamma_{A_{1}}A_{1}+\gamma_{A_{2}}A_{2}+\gamma_{Z_{1}}Z_{1}+\gamma_{Z_{2}}Z_{2}+\gamma_{Z_{3}}Z_{3}+\gamma_{M^{Y}}M^{Y}+\gamma_{M^{W}}M^{W}+\gamma_{M^{D}}M^{D},\varepsilon^{2} \right),$$

$$Pr\left( D=1 \right)\sim expit\left\{ \kappa_{0}+\kappa_{Y}Y+\kappa_{X}X+\kappa_{D}D+\kappa_{A_{1}}A_{1}+\kappa_{A_{2}}A_{2}+\kappa_{Z_{1}}Z_{1}+\kappa_{Z_{2}}Z_{2}+\kappa_{Z_{3}}Z_{3}+\kappa_{M^{Y}}M^{Y}+\kappa_{M^{X}}M^{X}+\kappa_{M^{D}}M^{D} \right\},$$

$$Pr\left( Y=1 \right)\sim expit\left\{ \lambda_{0}+\lambda_{Y}Y+\lambda_{X}X+\lambda_{D}D+\lambda_{A_{1}}A_{1}+\lambda_{A_{2}}A_{2}+\lambda_{Z_{1}}Z_{1}+\lambda_{Z_{2}}Z_{2}+\lambda_{Z_{3}}Z_{3}+\delta^{NARFCS}M^{Y}+\lambda_{M^{X}}M^{X}+\lambda_{M^{W}}M^{W}+\lambda_{M^{D}}M^{D} \right\}.$$

#### **1.5 Multiple imputation assuming missing at random**

We applied fully conditional specification using the following regressions:

$$Pr\left( W=1 \right)\sim expit\left\{ \iota_{0}+\iota_{Y}Y+\iota_{X}X+\iota_{D}D+\iota_{A_{1}}A_{1}+\iota_{A_{2}}A_{2}+\iota_{Z_{1}}Z_{1}+\iota_{Z_{2}}Z_{2}+\iota_{Z_{3}}Z_{3} \right\},$$

$X\sim N\left( \rho_{0}+\rho_{Y}Y+\rho_{W}W+\rho_{D}D+\rho_{A_{1}}A_{1}+\rho_{A_{2}}A_{2}+\rho_{Z_{1}}Z_{1}+\rho_{Z_{2}}Z_{2}+\rho_{Z_{3}}Z_{3},\varsigma^{2} \right),$

$$Pr\left( D=1 \right)\sim expit\left\{ \phi_{0}+\phi_{Y}Y+\phi_{X}X+\phi_{W}W+\phi_{A_{1}}A_{1}+\phi_{A_{2}}A_{2}+\phi_{Z_{1}}Z_{1}+\phi_{Z_{2}}Z_{2}+\phi_{Z_{3}}Z_{3} \right\},$$

$$Pr\left( Y=1 \right)\sim expit\left\{ \tau_{0}+\tau_{X}X+\tau_{W}W+\tau_{D}D+\tau_{A_{1}}A_{1}+\tau_{A_{2}}A_{2}+\tau_{Z_{1}}Z_{1}+\tau_{Z_{2}}Z_{2}+\tau_{Z_{3}}Z_{3} \right\}.$$

#### **1.6 Inverse probability weighting assuming missing at random**

The combination of missing data for $Y$ and for $X,W,A_{1},A_{2}$ and $D$ complicated the application of inverse probability weighting (IPW) because (i) there were multiple missingness mechanisms, and (ii) the factors influencing missingness were partially observed and the pattern of missingness was nonmonotone. In the absence of a correct weighting model applied to all participants, we considered IPW using three weighting models as summarised in supplementary table 2, along with the definition of the unstabilised and stabilised weights. All three weighting models were logistic regression models and assumed data were missing at random (MAR).

|  |  |  | **Unstabilised weights** | |  | **Stabilised weights** | |
| --- | --- | --- | --- | --- | --- | --- | --- |
| **Weighting model** | **Restricted to**  **participants** | **Dependent variable** | **Independent variables** | **Fitted probabilities to generate weights** |  | **Independent variables** | **Fitted probabilities to generate weights** |
| Main model | With observed values for $X$ and $D$ | $R^{main}=1$ when $Y$ and $W$ were observed; $R^{main}=0$ otherwise | $X,A_{1},A_{2},D$ | $\frac{1}{Pr\left( R^{main}=1 \vert X,A_{1},A_{2},D \right)}$ |  | $X$ | $\frac{Pr\left( R^{main}=1 \vert X \right)}{Pr\left( R^{main}=1 \vert X,A_{1},A_{2},D \right)}$ |
| Alternative model 1 | With observed values for $X,W$ and $D$ | $R^{alt1}=1$ when $Y$ was observed; $R^{alt1}=0$ otherwise | $X,A_{1},A_{2},D$ | $\frac{1}{Pr\left( R^{alt1}=1 \vert X,A_{1},A_{2},D \right)}$ |  | $X$ | $\frac{Pr\left( R^{alt1}=1 \vert X \right)}{Pr\left( R^{alt1}=1 \vert X,A_{1},A_{2},D \right)}$ |
| Alternative model 2 | With observed values for $X$ and $D$ | $R^{alt2}=1$ when $Y$ and $W$ were observed; $R^{alt2}=0$ otherwise | $X,A_{1},A_{2},D,$ $Z_{1},Z_{2},Z_{3}$ | $\frac{1}{Pr\left( R^{alt2}=1 \vert X,A_{1},A_{2},D,Z_{1},Z_{2},Z_{3} \right)}$ |  | $X,Z_{1},Z_{2},Z_{3}$ | $\frac{Pr\left( R^{alt2}=1 \vert X,Z_{1},Z_{2},Z_{3} \right)}{Pr\left( R^{alt2}=1 \vert X,A_{1},A_{2},D,Z_{1},Z_{2},Z_{3} \right)}$ |

**Supplementary table 2**: Summary of the weighting models considered for inverse probability weighting assuming missing at random

### **2 Methods: additional details for the motivating example**

#### **2.1 Summary statistics and investigations of missingness mechanisms**

**Supplementary table 3:** Missing data patterns of the substantive analysis variables: outcome (SARS-CoV-2 infection), exposure (body mass index), and confounders (smoker and degree) for 420,847 participants of UK Biobank Study. ✓ observed, $\times$ denotes missing, and ✓/ $\times$ denotes some observed and some missing.

| **Pattern** | **Outcome** | **Exposure** | **Confounders** | **Number of participants (%)** |
| --- | --- | --- | --- | --- |
| 1 | ✓ | ✓ | ✓ | 4,610 (1.10%) |
| 2 | ✓ | ✓ | ✓/ $\times$ | 136 (0.0323%) |
| 3 | ✓ | $\times$ | ✓ | 50 (0.0119%) |
| 4 | ✓ | $\times$ | ✓/ $\times$ | 7 (0.00166%) |
| 5 | $\times$ | ✓ | ✓ | 405,174 (96.3%) |
| 6 | $\times$ | ✓ | ✓/ $\times$ | 8,492 (2.02%) |
| 7 | $\times$ | $\times$ | ✓ | 1,936 (0.460%) |
| 8 | $\times$ | $\times$ | ✓/ $\times$ | 442 (0.105%) |

Using the observed data, we investigated whether the chance of a missing outcome depended on (i) the substantive analysis covariates (body mass index (BMI), age (age), sex, degree, and current smoking status) and (ii) the auxiliaries (asthma, diabetes, and hypertension diagnoses) after conditioning on the substantive analysis covariates. We used a logistic regression model, which we refer to as the “missingness model”, in which the dependent variable was a binary variable,$M^{outcome}$, indicating whether the individual had an observed or missing value for outcome, SARS-CoV-2 infection ($M^{outcome}=0$ for observed SARS-CoV-2 infection and $M^{outcome}=1$ otherwise). We fitted two missingness models to the data in which the independent variables were (i) the substantive analysis covariates and (ii) the substantive analysis covariates and the auxiliary variables.

A similar approach was used to identify predictors of the missingness mechanism for a

missing covariate (BMI, degree, or smoker) by setting the dependent variable of the

missingness model to be a binary variable indicating whether an individual had complete data on all substantive analysis covariates ($M^{covariate}=0$) or missing data on at least one covariate ($M^{covariate}=1$). Here, the independent variables of these missingness models were age, sex, and infection with SARS-CoV-2 with or without the auxiliary variables.

Supplementary table 4 shows that the chance of having a missing outcome was associated with the observed values of the exposure, the confounders, and the auxiliary variables.

Additionally, supplementary table 5 shows that missingness of the covariates depended on the auxiliary variables (i.e., participants with a co-morbidity were more likely to have missing data on current smoking status, BMI, and highest educational attainment than participants without a co-morbidity). Unsurprisingly, given that SARS-CoV-2 infection occurred more than a decade after measurement of the (baseline) covariates, there is no evidence that missingness of these variables depends on SARS-CoV-2 infection status (after conditioning on fully observed baseline variables). Note that, we cannot use these investigations based on the observed data to inform us whether missingness of the outcome or covariates depended on the missing values of the outcome or covariates. These decisions were made based on subject-matter knowledge as described in the main text.

**Supplementary table 4:** Associations with the odds of a missing outcome among 409,784 participants with observed data on body mass index (BMI), degree and smoker. UK Biobank Study.

|  | **Odds ratio (95% confidence interval)** | |
| --- | --- | --- |
|  | **Missingness model 1** | **Missingness model 2** |
| BMI^std^ | 0.827 (0.805, 0.849) | 0.962 (0.935, 0.990) |
| age^std^ | 0.916 (0.889, 0.944) | 1.03 (0.997, 1.06) |
| Sex | 0.892 (0.842, 0.946) | 0.970 (0.914, 1.03) |
| Degree | 1.16 (1.08, 1.23) | 1.09 (1.02, 1.16) |
| Smoker | 0.725 (0.664, 0.792) | 0.832 (0.762, 0.910) |
| Asthma | - | 0.368 (0.347, 0.391) |
| Diabetes | - | 0.579 (0.533, 0.628) |
| Hypertension | - | 0.635 (0.594, 0.678) |

age^std^: Age standardised to have mean 0 and standard deviation of 1; BMI^std^: BMI standardised to have mean 0 and standard deviation of 1.

**Supplementary table 5:** Associations with the odds of a missing covariate (body mass index, smoker, or degree) among 4,803 participants with observed data on SARS-CoV-2 infection. Data from the UK Biobank Study.

|  | **Odds ratio (95% confidence interval)** | |
| --- | --- | --- |
|  | **Missingness model 1** | **Missingness model 2** |
| SARS-Cov-2 infection | 1.07 (0.775, 1.46) | 1.04 (0.752, 1.42) |
| age^std^ | 1.14 (0.991, 1.31) | 0.991 (0 .855, 1.15) |
| Sex | 1.22 (0.910, 1.64) | 1.09 (0.814, 1.47) |
| Asthma | - | 1.32 (0.978, 1.80) |
| Diabetes | - | 1.51 (1.08, 2.10) |
| Hypertension | - | 1.73 (1.21, 2.49) |

age^std^: Age standardised to have mean 0 and standard deviation of 1.

**Supplementary table 6:** Characteristics of the UK Biobank participants among the 420,847 eligible for analysis and the 409,784 with complete data on exposure (body mass index) and confounders (age, sex, degree, and current smoker)

| **Characteristic** | **N (%) or mean (SD)** | |
| --- | --- | --- |
|  | **Eligible for analysis**  **N=** **420,847** | **Complete data on exposure & confounders N=409,784** |
| Tested for SARS-CoV-2 |  |  |
| No | 416,044 (98.9%) | 405,174 (98.9%) |
| Yes | 4,803 (1.14%) | 4,610 (1.12%) |
| Missing | 0 (0%) | 0 (0%) |
| Positive for SARS-CoV-2 |  |  |
| No | 3,428 (0.814%) | 3,293 (0.804 %) |
| Yes | 1,375 (0.327%) | 1,317 (0.321%) |
| Missing | 416,044 (98.9%) | 405,174 (98.9%) |
| Body Mass Index (kg/m^2^) |  |  |
|  | 27.4 (4.75) | 27.3 (4.75) |
| Missing | 2,435 (0.579%) | 0 (0%) |
| Age at baseline (years) |  |  |
|  | 56.3 (8.09) | 56.3 (8.08) |
| Missing | 0 (0%) | 0 (0%) |
| Sex |  |  |
| Women | 231,877 (55.1%) | 226,030 (55.2%) |
| Men | 188,970 (44.9%) | 183,754 (44.8%) |
| Missing | 0 (0%) | 0 (0%) |
| Degree-level education |  |  |
| No | 275,490 (66.7%) | 272,976 (66.6%) |
| Yes | 137,661 (33.3%) | 136,808 (33.4%) |
| Missing | 7,696 (1.83%) | 0 (0%) |
| Current smoker |  |  |
| No | 377,132 (89.6%) | 369,614 (90.2%) |
| Yes | 41,370 (9.83%) | 40,170 (9.80%) |
| Missing | 2,345 (0.56%) | 0 (0%) |
| Asthma |  |  |
| No | 317,543 (75.5%) | 309,669 (75.6%) |
| Yes | 103,3.04 (24.5%) | 100,115 (24.4%) |
| Missing | 0 (0%) | 0 (0%) |
| Diabetes |  |  |
| No | 383,973 (91.2%) | 374,597 (91.4%) |
| Yes | 36,874 (8.76%) | 35,187 (8.59%) |
| Missing | 0 (0%) | 0 (0%) |
| Hypertension |  |  |
| No | 262,890 (62.5%) | 256,983 (62.7%) |
| Yes | 157,957 (37.5%) | 152,801 (37.3%) |
| Missing | 0 (0%) | 0 (0%) |

#### **2.2 Details of missing data methods applied to UK Biobank study**

**Supplementary table 7**: Specifications of the imputation, weighting and bias models applied to data from the UK Biobank study

| **Method** | **Model** | **Type of regression** | **Dependent variable** | **Independent variables** | **Prior distribution** |
| --- | --- | --- | --- | --- | --- |
| MI | imputation model assuming MAR^a^ | Logistic | SARS-CoV-2 | sex, age^std^, degree, smoker, BMI^std^, asthma, diabetes, hypertension | $p\left( \nu\right)\propto1$ for coefficients $\nu$ |
| IPW | weighting model assuming MAR | Logistic | tested (tested=1 when SARS-Cov-2 is observed; tested=0 otherwise) | sex, age^std^, degree, smoker, BMI^std^, asthma, diabetes, hypertension | Not applicable |
| Monte Carlo NARFCS | pattern-mixture imputation model | Logistic | SARS-CoV-2 | sex, age^std^, degree, smoker, BMI^std^, asthma, diabetes, hypertension, $M^{SARSCoV2}$ ($M^{SARSCoV2}=1$ when SARS-CoV-2 is missing and $M^{SARSCoV2}=0$ otherwise) | $p\left( \delta^{NARFCS} \right)\sim N\left( -2.60,{0.220}^{2} \right)$ and $p\left( \nu\right)\propto1$ for remaining coefficients $\nu$ |
|  |  |  | Coefficient of $M^{SARSCoV2}$ is bias parameter $\delta^{NARFCS}$ | |  |
| Bayesian SM | sequential, selection model | Logistic | SARS-CoV-2 | sex, age^std^, degree, smoker, BMI^std^, | $p\left( \delta^{SM} \right)\sim N\left( 2.60,{0.220}^{2} \right)$ and $N\left( 0,100 \right)$ for all remaining regression coefficients |
|  |  | Logistic | asthma | sex, age^std^, degree, smoker, BMI^std^, SARS-CoV-2 |  |
|  |  | Logistic | diabetes | sex, age^std^, degree, smoker, BMI^std^, SARS-CoV-2, asthma |  |
|  |  | Logistic | hypertension | sex, age^std^, degree, smoker, BMI^std^, SARS-CoV-2, asthma, diabetes |  |
|  |  | Logistic | tested | sex, age^std^, degree, smoker, BMI^std^, SARS-CoV-2, asthma, diabetes, hypertension |  |
|  |  |  | Coefficient of SARS-CoV-2 is bias parameter $\delta^{SM}$ | |  |

a: Missing At Random; age^std^: Age standardised to have mean 0 and standard deviation of 1; BMI^std^: BMI standardised to have mean 0 and standard deviation of 1.

Substantive analysis of interest

$$Pr\left( SARSCoV2=1 \right)\sim expit\left\{ \beta_{cons}+\beta_{{BMI}^{std}}{BMI}^{std}+\beta_{{age}^{std}}{age}^{std}+\beta_{sex}sex+\beta_{smoker}smoker+\beta_{degree}degree \right\}$$

Imputation model of MI

$$Pr\left( SARSCoV2=1 \right)\sim expit\left\{ \tau_{cons}+\tau_{{BMI}^{std}}{BMI}^{std}+\tau_{{age}^{std}}{age}^{std}+\tau_{sex}sex+\tau_{smoker}smoker+\tau_{degree}degree+\tau_{asthma}asthma+\tau_{diabetes}diabetes+\tau_{hyper}hyperten \right\}$$

Weighting model of IPW

$$Pr\left( tested=1 \right)\sim expit\left\{ \varpi_{cons}+\varpi_{{BMI}^{std}}{BMI}^{std}+\varpi_{{age}^{std}}{age}^{std}+\varpi_{sex}sex+\varpi_{smoker}smoker+\varpi_{degree}degree+\varpi_{asthma}asthma+\varpi_{diabetes}diabetes+\varpi_{hyper}hyperten \right\}$$

Bias model of Monte Carlo NARFCS

$$Pr\left( SARSCoV2=1 \right)\sim expit\left\{ \lambda_{0}+\lambda_{{BMI}^{std}}{BMI}^{std}+\lambda_{{age}^{std}}{age}^{std}+\lambda_{sex}sex+\lambda_{smoker}smoker+\lambda_{degree}degree+\lambda_{asthma}asthma+\lambda_{diabetes}diabetes+\lambda_{hyper}hyperten+\delta^{NARFCS}M^{SARSCoV2} \right\}$$

Bias model of Bayesian SM

$$Pr\left( SARSCoV2=1 \right)\sim expit\left\{ \beta_{cons}+\beta_{{BMI}^{std}}{BMI}^{std}+\beta_{{age}^{std}}{age}^{std}+\beta_{sex}sex+\beta_{smoker}smoker+\beta_{degree}degree \right\},$$

$$Pr\left( asthma=1 \right)\sim expit\left\{ \theta_{cons}+\theta_{{BMI}^{std}}{BMI}^{std}+\theta_{{age}^{std}}{age}^{std}+\theta_{sex}sex+\theta_{smoker}smoker+\theta_{degree}degree+\theta_{SARSCoV2}SARSCoV2 \right\},$$

$$Pr\left( diabetes=1 \right)\sim expit\left\{ o_{cons}+o_{{BMI}^{std}}{BMI}^{std}+o_{{age}^{std}}{age}^{std}+o_{sex}sex+o_{smoker}smoker+o_{degree}degree+o_{SARSCoV2}SARSCoV2+o_{asthma}asthma \right\},$$

$$Pr\left( hyperten=1 \right)\sim expit\left\{ \omega_{cons}+\omega_{{BMI}^{std}}{BMI}^{std}+\omega_{{age}^{std}}{age}^{std}+\omega_{sex}sex+\omega_{smoker}smoker+\omega_{degree}degree+\omega_{SARSCoV2}SARSCoV2+\omega_{asthma}asthma+\omega_{diabetes}diabetes \right\},$$

$$Pr\left( tested=1 \right)\sim expit\left\{ \psi_{cons}+\psi_{{BMI}^{std}}{BMI}^{std}+\psi_{{age}^{std}}{age}^{std}+\psi_{sex}sex+\psi_{smoker}smoker+\psi_{degree}degree+\delta^{SM}SARSCoV2+\psi_{asthma}asthma+\psi_{diabetes}diabetes+\psi_{hyper}hyperten \right\}.$$

##### **2.2.1 Deriving values of the prior hyperparameters for the bias parameter**

First, consider bias parameter $\delta^{NARFCS}$ which denotes the difference in the log-odds of being infected with SARS-CoV-2 between those with a missing and observed value for SARS-CoV-2, conditional on sex, age, BMI, degree, smoker, asthma, diabetes, and hypertension.

In the absence of external information about the likely values of $\delta^{NARFCS}$, we used results from a published study reporting SARS-CoV-2 antibody prevalence during the time period of interest. We first describe the algorithm used to convert information about marginal prevalences to information about $\delta^{NARFCS}$, and then we describe how we derived values for the hyperparameters of prior $p\left( \delta^{NARFCS} \right)\sim N\left( \mu^{NARFCS},\sigma^{NARFCS} \right)$.

For a given prevalence of SARS-CoV-2 infection, $\pi^{target}$, we computed a calibrated value of $\delta^{NARFCS}$ using the following algorithm from Tompsett et al (1):

A. Specify a tolerance for $\pi^{target}$ (e.g., $\pm0.02$).
B. Specify a range of test values for $\delta^{NARFCS}$ and take points in regular intervals over this range.
C. For each test value, $\hat{\delta}^{NARFCS(TV)}$, specified in step B, repeat the following steps (using the same random seed number each time):
   i. multiply impute variable SARS-CoV-2 (5 imputations) using the NARFCS bias model with $\delta^{NARFCS}=\hat{\delta}^{NARFCS(TV)}$.
   ii. using the multiply imputed data from step (i), estimate the prevalence of infection separately in each imputed dataset and combine the results using Rubin’s rules to give point estimate $\hat{\pi}^{\delta^{NARFCS(TV)}}$
D. Among the prevalence point estimates of step C, determine the two values of $\hat{\pi}^{\delta^{NARFCS(TV)}}$ that are closet to $\pi^{target}$ (i.e., $\hat{\pi}_{lower}^{NARFCS(TV)}\leq\pi^{target}\leq\hat{\pi}_{upper}^{NARFCS(TV)}$) and their corresponding values for $\delta^{NARFCS}$ (i.e., $\delta_{lower}^{NARFCS(TV)}$ and $\delta_{upper}^{NARFCS(TV)})$. If $\hat{\pi}_{lower}^{NARFCS(TV)}$ or $\hat{\pi}_{upper}^{NARFCS(TV)}$ is within the specified tolerance, then choose the value closest to $\pi^{target}$ and exit the algorithm. Otherwise, return to step B setting the range of test values to be between $\delta_{lower}^{NARFCS(TV)}$ and $\delta_{upper}^{NARFCS(TV)}$.

For value $\hat{\pi}_{lower}^{NARFCS(TV)}$ (or $\hat{\pi}_{upper}^{NARFCS(TV)}$) closest to $\pi^{target}$, the corresponding value $\delta_{lower}^{NARFCS(TV)}$ (or $\delta_{upper}^{NARFCS(TV)}$) becomes the value of $\delta^{NARFCS}$ calibrated to $\pi^{target}$.

Among 65–74-year-olds of the REal-time Assessment of Community Transmission-2 (REACT-2) national study (2), SARS-CoV-2 antibody prevalence was estimated to be 3.2% [95% confidence interval (CI) 2.8–3.6%] by mid-July 2020. Given the similarities between the REACT-2 study and our UK Biobank (UKB) study (i.e., age range, geographical location, and time period), we initially considered the prevalence of SARS-CoV-2 infection in our UKB study could plausibly be 3.2% [95% CI 2.2–4.2%], allowing for additional uncertainty about the unknown prevalence of infection in our UKB study.

Using a tolerance of $\pm0.02$ (in step A) and test values for $\delta^{NARFCS(TV)}$ of between $-4$ and $-1$ in regular intervals of $0.25$ (in step B), the value of $\delta^{NARFCS}$ calibrated to 3.2% was -2.56 and the calibrated values for interval 2.2% to 4.2% were -3.00 and -2.25, respectively. Initially, we set $\mu^{NARFCS}=-2.56$ and then derived the value for $\sigma^{NARFCS}$ as follows:

I. Calculated the standard deviation of the Normal distribution $N\left( -2.56,\sigma_{-3.00}^{NARFCS} \right)$ such that the 2.5^th^ percentile was -3.00

$$\sqrt{\sigma_{-3.00}^{NARFCS}}=\frac{\left( -2.56+3.00 \right)}{1.96}=0.2245$$

II. Calculated the standard deviation of the Normal distribution $N\left( -2.56,\sigma_{-2.25}^{NARFCS} \right)$ such that the 97.5^th^ percentile was -2.25

$$\sqrt{\sigma_{-2.25}^{NARFCS}}=\frac{\left( -2.25+2.6 \right)}{1.96}=0.1582$$

III. We chose the larger standard error rounded to two significant figures.

Finally, we decided to round the mean value to two significant figures so that our final prior was $p\left( \delta^{NARFCS} \right)\sim N\left( -2.6,{0.22}^{2} \right)$.

Bias parameter $\delta^{SM}$ denotes the difference in the log-odds of being *tested* with SARS-CoV-2 between those infected and not infected with SARS-CoV-2, conditional on sex, age, degree, smoker, BMI, SARS-CoV-2, asthma, diabetes, and hypertension. For Bayesian SM we used prior $p\left( \delta^{SM} \right)\sim N\left( \mu^{SM},\sigma^{SM} \right)$ for $\delta^{SM}$. We applied the above algorithm to calculate values of $\delta^{SM}$ calibrated to a given prevalence of SARS-CoV-2 infection, replacing the application of NARFCS in step C with Bayesian SM. Our investigations showed that the calibrated values of $\delta^{SM}$ and $\delta^{NARFCS}$ were very similar. Therefore, we set $\mu^{SM}=-\mu^{NARFCS}=2.6$, backed-up by the following reasoning: (i) the symmetrical property of the odds ratio (i.e., the same odds ratio is obtained from the logistic regression of infection status on tested and the logistic regression of tested on infection status). (ii) variable tested (1 if tested, 0 if not tested) used by Bayesian SM is coded in the opposite direction to the missingness indicator for SARS-CoV-2 (1 if missing SARS-CoV-2 (i.e., not tested), 0 otherwise (i.e., tested)) used by Monte Carlo NARFCS, and (iii) $\delta^{SM}$ and $\delta^{NARFCS}$ describe the association between missingness and SARS-CoV-2 infection conditional on the same set of variables. We set $\sigma^{SM}=\sigma^{NARFCS}={0.22}^{2}$, and so the final choice for the prior was $p\left( \delta^{SM} \right)\sim N\left( 2.6,{0.22}^{2} \right)$.

### **3 Additional simulation study results**

#### **3.1 Main results for selection model data generating model**

**Supplementary table 8:** Summary of results when $\beta_{X}=\ln3$ and simulation using selection model data generating model: number of simulated datasets (No. Sim.), bias and empirical standard error (Emp SE) of estimate $\hat{\beta}_{X}$, mean of model-based SE of $\hat{\beta}_{X}$, coverage of 95% confidence interval for $\hat{\beta}_{X}$. [95% Monte Carlo interval].

|  | **No.**  **Sim.** | **Bias** | **Emp. SE** | **Mean SE^$^** | **Coverage %** |
| --- | --- | --- | --- | --- | --- |
| Full data analysis | 1000 | 0.000844  [-0.000173, 0.00186] | 0.0164  [0.0157, 0.0171] | 0.0164  [0.0164, 0.0165] | 94.5  [93.1, 95.9] |
| Complete case analysis | 1000 | -0.139  [-0.144, -0.135] | 0.0710  [0.0678, 0.0741] | 0.0690  [0.0689, 0.0692] | 47.8  [44.7, 50.9] |
| MI | 1000 | -0.223 [-0.227, -0.218] | 0.0688 [0.0658, 0.0719] | 0.0666 [0.0661, 0.0670] | 9.50 [7.68, 11.3] |
| IPW; main model; unstabilised weights | 1000 | -0.141 [-0.150, -0.132] | 0.145 [0.139, 0.152] | 0.135 [0.134, 0.136] | 79.5 [77.0, 82.0] |
| IPW; main model; stabilised weights | 1000 | -0.153 [-0.159, -0.147] | 0.0942 [0.0900, 0.0983] | 0.0930 [0.0925, 0.0934] | 60.4 [57.4, 63.4] |
| Monte Carlo NARFCS^&^ with very informative prior | 1000 | -0.00121 [-0.00240, -0.0000119] | 0.0193 [0.0184, 0.0201] | 0.0226 [0.0225, 0.0227] | 95.9 [94.7, 97.1] |
| Monte Carlo NARFCS^&^ with informative prior | 1000 | -0.00358 [-0.00479, -0.00237] | 0.0195 [0.0186, 0.0203] | 0.0385 [0.0380, 0.0391] | 97.6 [96.7, 98.5] |
| Monte Carlo NARFCS^&^ with vague prior | 1000 | -0.215 [-0.219, -0.211] | 0.0678 [0.0648, 0.0707] | 0.489 [0.488, 0.490] | 94.4 [93.0, 95.8] |
| Bayesian SM with very informative prior | 927 | -0.00971  [-0.0137, -0.00570] | 0.0624  [0.0596, 0.653] | 0.0269  [0.0244, 0.0295] | 92.3  [90.6, 94.1] |
| Bayesian SM with informative prior | 928 | -0.0330  [-0.0406, -0.0254] | 0.118  [0.113, 0.124] | 0.0354  [0.0322, 0.0386] | 87.8  [85.7, 89.9] |
| Bayesian SM with vague | 926 | -0.0657  [-0.0810, -0.0503] | 0.238  [0.228, 0.249] | 0.0719  [0.0634, 0.0804] | 87.4  [85.2, 89.5] |

$ For Bayesian SM: standard deviation of the posterior distribution of $\hat{\beta}_{X}$, and for Monte Carlo NARFCS: standard deviation of the frequency distribution of $\hat{\beta}_{X}^{\delta^{NARFCS(1)}},\hat{\beta}_{X}^{\delta^{NARFCS(2)}},\cdots,\hat{\beta}_{X}^{\delta^{NARFCS(S)}}$. & Monte Carlo NARFCS using 10,000 Monte Carlo steps with single imputation.

**Supplementary table 9:** Summary of results when $\beta_{X}=0$ and simulation using selection model data generating model: number of simulated datasets (No. Sim.), bias and empirical standard error (Emp SE) of estimate $\hat{\beta}_{X}$, mean of model-based SE of $\hat{\beta}_{X}$, coverage of 95% confidence interval for $\hat{\beta}_{X}$. [95% Monte Carlo interval].

|  | **No.**  **Sim.** | **Bias** | **Emp. SE** | **Mean SE^$^** | **Coverage %** |
| --- | --- | --- | --- | --- | --- |
| Full data analysis | 1000 | -0.0000897  [-0.00101, 0.000826] | 0.0148  [0.0141, 0.0154] | 0.0145  [0.0145, 0.0145] | 94.9  [93.5, 96.3] |
| Complete case analysis | 1000 | -0.139  [-0.143, -0.136] | 0.0544  [0.0520, 0.0567] | 0.0540  [0.0540, 0.0541] | 26.9  [24.2, 29.6] |
| MI | 1000 | -0.132 [-0.135, -0.129] | 0.0517 [0.0494, 0.0540] | 0.0522 [0.0519, 0.0525] | 28.4 [25.6, 31.2] |
| IPW; main model; unstabilised weights | 1000 | -0.131 [-0.134, -0.127] | 0.0557 [0.0533, 0.0582] | 0.0565 [0.0563, 0.0566] | 36.5 [33.5, 39.5] |
| IPW; main model; stabilised weights | 1000 | -0.132 [-0.135, -0.128] | 0.0555 [0.0531, 0.0580] | 0.0564 [0.0562, 0.0565] | 35.6 [32.6, 38.6] |
| Monte Carlo NARFCS^&^ with very informative prior | 1000 | -0.00206 [-0.00314, -0.000975] | 0.0175 [0.0167, 0.0182] | 0.0231  [0.0229, 0.0233] | 97.3 [96.3, 98.3] |
| Monte Carlo NARFCS^&^ with informative prior | 1000 | -0.00379 [-0.00488, -0.00271] | 0.0175 [0.0168, 0.0183] | 0.0372  [0.0366, 0.0377] | 98.0 [97.1, 98.9] |
| Monte Carlo NARFCS^&^ with vague prior | 1000 | -0.106 [-0.109, -0.103] | 0.0470 [0.0449, 0.0490] | 0.0822  [0.0811, 0.0832] | 96.9 [95.8, 98.0] |
| Bayesian SM with very informative prior | 928 | -0.00689  [-0.00821, -0.00557] | 0.0295  [0.0196, 0.0214] | 0.0331  [0.0323, 0.0339] | 98.8  [98.1, 99.5] |
| Bayesian SM with informative prior | 928 | -0.0248  [-0.0272, -0.0224] | 0.0376  [0.0359, 0.0393] | 0.0431  [0.0422, 0.0439] | 93.4  [91.8, 95.0] |
| Bayesian SM with vague | 929 | -0.0418  [-0.0448, -0.0387] | 0.0474  [0.0452, 0.0495] | 0.0449  [0.0442, 0.0457] | 82.6  [80.1, 85.0] |

$ For Bayesian SM: standard deviation of the posterior distribution of $\hat{\beta}_{X}$, and for Monte Carlo NARFCS: standard deviation of the frequency distribution of $\hat{\beta}_{X}^{\delta^{NARFCS(1)}},\hat{\beta}_{X}^{\delta^{NARFCS(2)}},\cdots,\hat{\beta}_{X}^{\delta^{NARFCS(S)}}$. & Monte Carlo NARFCS using 10,000 Monte Carlo steps with single imputation.

**Supplementary table 10:** Summary of the results of inverse probability weighting assuming missing at random using the main and alternative weighting models when $\beta_{X}=\ln3$ and $\beta_{X}=0$: Bias and empirical standard error (Emp SE) of estimate $\hat{\beta}_{X}$, mean of the model-based SE of $\hat{\beta}_{X}$, coverage of the 95% confidence interval for $\hat{\beta}_{X}$. [95% Monte Carlo interval]. Simulation using selection model data generating model. Results based on 1000 simulated datasets.

|  | **True value of** $\beta_{X}$ | **Bias** | **Emp. SE** | **Mean SE** | **Coverage %** |
| --- | --- | --- | --- | --- | --- |
| Main model; unstabilised weights | $\ln3$ | -0.141 [-0.150, -0.132] | 0.145 [0.139, 0.152] | 0.135 [0.134, 0.136] | 79.5 [77.0, 82.0] |
| Alternative weighting model 1; unstabilised weights | $\ln3$ | -0.141  [-0.151, -0.132] | 0.148 [0.142, 0.155] | 0.137 [0.136, 0.138] | 79.7  [77.2, 82.2] |
| Alternative weighting model 2; unstabilised weights | $\ln3$ | -0.147  [-0.156, -0.138] | 0.146 [0.140, 0.153] | 0.136 [0.135, 0.137] | 78.9  [76.4, 81.4] |
| Main model; stabilised weights | $\ln3$ | -0.153 [-0.159, -0.147] | 0.0942 [0.0900, 0.0983] | 0.0930 [0.0925, 0.0934] | 60.4 [57.4, 63.4] |
| Alternative weighting model 1; stabilised weights | $\ln3$ | -0.154 [-0.160, -0.148] | 0.0947 [0.0906, 0.0989] | 0.0935 [0.0931, 0.0940] | 60.6 [57.6, 63.6] |
| Alternative weighting model 2; stabilised weights | $\ln3$ | -0.157 [-0.163, -0.151] | 0.0948 [0.0906, 0.0989] | 0.0939 [0.0935, 0.0944] | 60.0 [57.0, 63.0] |
| Main model; unstabilised weights | 0 | -0.131 [-0.134, -0.127] | 0.0557 [0.0533, 0.0582] | 0.0565 [0.0563, 0.0566] | 36.5 [33.5, 39.5] |
| Alternative weighting model 1; unstabilised weights | $0$ | -0.129  [-0.133, -0.126] | 0.0558 [0.0534, 0.0583] | 0.0566 [0.0564, 0.0567] | 38.0  [35.0, 41.0] |
| Alternative weighting model 2; unstabilised weights | $0$ | -0.131  [-0.134, -0.127] | 0.0569 [0.0544, 0.0594] | 0.0574 [0.0572, 0.0575] | 39.0  [36.0, 42.0] |
| IPW; main model; stabilised weights | 0 | -0.132 [-0.135, -0.128] | 0.0555 [0.0531, 0.0580] | 0.0564 [0.0562, 0.0565] | 35.6 [32.6, 38.6] |
| Alternative weighting model 1; stabilised weights | $0$ | -0.130 [-0.134, -0.127] | 0.0556 [0.0532, 0.0581] | 0.0564 [0.0563, 0.0566] | 36.6 [33.6, 39.6] |
| Alternative weighting model 2; stabilised weights | $0$ | -0.134 [-0.137, -0.131] | 0.0553 [0.0529, 0.0578] | 0.0562 [0.0562, 0.0563] | 33.6 [30.7, 36.5] |

#### **3.2 Multiple imputation assuming missing at random; imputing with and**

#### **without auxiliary information**

In this section we explore the larger than expected bias of multiple imputation (MI) in the not null scenario ($\beta_{X}=\ln3$) and report our investigations on the probable cause. As before, MI refers to multiple imputation under the MAR assumption.

Figure 1a (of the main manuscript) shows that in the not null scenario a CCA may be biased by missing data via direct edge Y🡪M^Y^ and pathways via auxiliary variables A and D (e.g., Y🡨A🡪 M^Y^ and Y🡨D🡪 M^Y^). For MI, with X, A and D included in the imputation model, bias due to missing data (excluding model misspecification) should only occur via direct edge Y🡪M^Y^ as the other pathways are closed by conditioning on X, A and D. Since the relationships of M^Y^ with Y, A and D were all in the same direction, we expected that closing the pathway via A and D would reduce the bias due to missing data compared to a CCA (3). However, for MI, with A and D included in the imputation model, the bias of the exposure effect was more than 50% larger than that of CCA (supplementary table 8).

To investigate, we simplified our simulation study such that only outcome Y was partially observed. The complete data were simulated using the SM data generating model. We considered three missingness mechanisms: Y missing completely at random (MCAR), Y MAR depending on X, A, and D, and Y missing not at random (MNAR) depending on Y, X, A, and D as depicted by the missingness directed acyclic graphs in supplementary figure 1. Missingness in Y was simulated using

$Pr\left( M^{Y}=1 \right)=expit\left\{ \psi_{0}^{SM}+\psi_{X}^{SM}X+\psi_{A_{1}}^{SM}A_{1}+\psi_{A_{2}}^{SM}A_{2}+\psi_{D}^{SM}D+\psi_{Y}^{SM}Y \right\}$,

where $A=\left( A_{1},A_{2} \right)$ and the parameter values are reported in supplementary table 11. With the exception of the missingness mechanism for Y and that X, W and D were fully observed in this simpler setting, all other settings were the same as per the main simulation study.

We compared a CCA, MI including A and D in the imputation model, MI excluding A and D from the imputation model, NARFCS including A and D, and NARFCS excluding A and D from the imputation model. Note that, in this simpler setting the imputation model of MI consisted of a single logistic regression with independent variables X, Z, W, and, when specified, A and D. Similarly, for NARFCS except the imputation models also included missingness indicator M^Y^. Also, we fitted NARFCS with its bias parameter fixed to its true value (i.e., did not implement NARFCS as a probabilistic bias analysis). Additionally, for the Y MNAR scenario, we repeated the simulation study using weaker relationships between A and Y and between D and Y (i.e., on the odds scale, the weaker relationships were 57% smaller than the original relationships).

Supplementary table 12 shows the results of the simplified simulation study. As expected, imputing using the same model as the substantive analysis (i.e., MI excluding A and D from the imputation model) gave similar levels of bias to a CCA. The results for Y MCAR and Y MAR show that including A and D in the imputation model did not induce bias due to misspecification of the imputation model. In fact, for Y MAR (depending on X, A and D), applying MI with A and D in the imputation model reduced the bias due to missing data (as expected). The magnitude of the bias of MI was only higher than that of CCA when conditioning on A and D, Y was MNAR and had not been accounted for, and A and D explained a substantial proportion of the variance of Y. Therefore, we concluded that variables A and D acted as amplifiers for bias due to Y MNAR (3).

Finally, for the null scenario of our main simulation study the relationships between Y and A and D were null. Therefore, MI including A and D did not amplify the bias due to data MNAR (see supplementary table 9).

|  | **Not null** $\left( \boldsymbol{\beta}_{\boldsymbol{X}}\boldsymbol{=}\ln\boldsymbol{3} \right)$ **scenario** | **Null** $\left( \boldsymbol{\beta}_{\boldsymbol{X}}\boldsymbol{=}\boldsymbol{0} \right)$ **scenario** |
| --- | --- | --- |
| $\boldsymbol{Y}$  **M**  **C**  **A**  **R** | 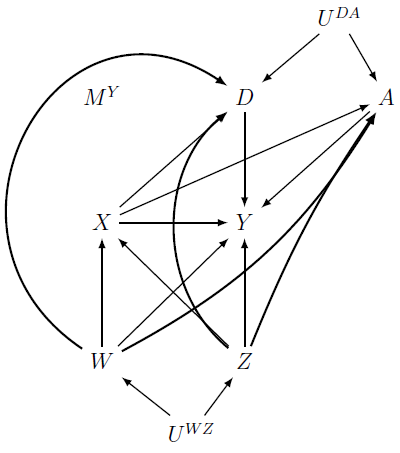 | 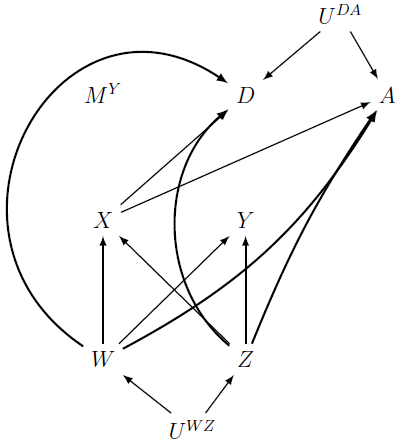 |
| $\boldsymbol{Y}$  **M**  **A**  **R** | 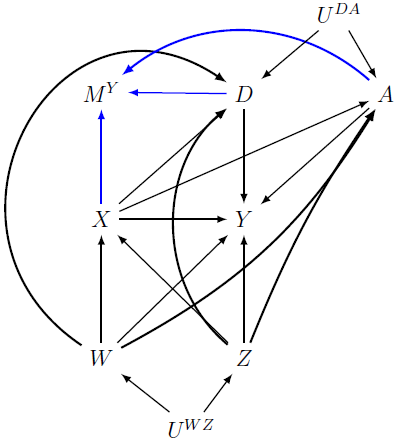 | 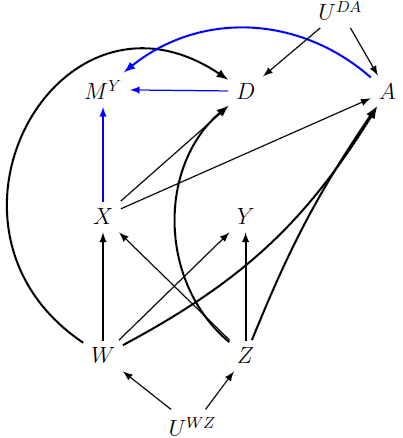 |
| $\boldsymbol{Y}$  **M**  **N**  **A**  **R** | 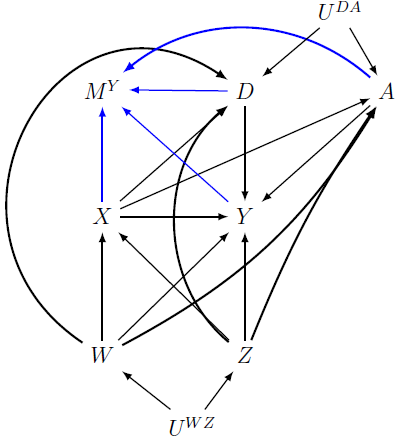 | 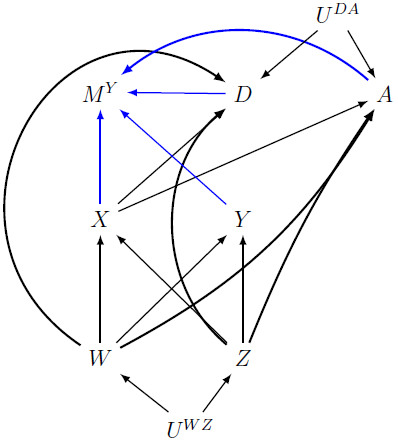 |

**Supplementary figure 1**: Missingness directed acyclic graphs (m-DAGs) of the simpler simulation study when the exposure effect, $\beta_{X}$, is not-null and null. Black and blue edges depict the relationships in the fully observed data and the outcome missingness mechanism, respectively.

**Supplementary table 11:** Parameter values of the missingness mechanism model for outcome Y in the simplified simulation study

| **Data model parameter** | **Y MCAR** | **Y MAR depending on X, A and D** | **Y MNAR depending on Y, X, A, and D** |
| --- | --- | --- | --- |
| $\psi_{0}^{SM}$ | 2.95 | 4.75 | 6.30 |
| $\psi_{X}^{SM}$ |  | -0.604 | -0.013 |
| $\psi_{A_{1}}^{SM}$ |  | -1.57 | -0.991 |
| $\psi_{A_{2}}^{SM}$ |  | -1.11 | -0.519 |
| $\psi_{D}^{SM}$ |  | -1.01 | -0.441 |
| $\psi_{Y}^{SM}$ |  |  | -7.85 |

**Supplementary table 12:** Summary of the simplified simulation results when $\beta_{X}=\ln3$: Bias of exposure effect estimate. [95% Monte Carlo interval].

| **Missingness mechanism** | **Method** | **Bias** |
| --- | --- | --- |
| $Y$ MCAR^a^ | Complete case analysis | 0.00164 [-0.00292, 0.00620] |
|  | MI excluding A and D | 0.00224 [-0.00274, 0.00723] |
|  | MI including A and D | -0.000646 [-0.00527, 0.00398] |
| $Y$ MAR^b^ | Complete case analysis | -0.130 [-0.133, -0.128] |
|  | MI excluding A and D | -0.131 [-0.133, -0.128] |
|  | MI including A and D | -0.00249 [-0.00542, 0.000438] |
| $Y$ MNAR^c^ | Complete case analysis | -0.136 [-0.140, -0.132] |
|  | MI excluding A and D | -0.135 [-0.139,-0.131] |
|  | MI including A and D | -0.208 [-0.213,-0.204] |
|  | NARFCS excluding A and D | 0.00325 [0.00204, 0.00447] |
|  | NARFCS including A and D | 0.00307 [0.00191, 0.00422] |
| $Y$ MNAR  Weaker $Y-A$, and $Y-D$ relationships | Complete case analysis | -0.140 [-0.144, -0.136] |
|  | MI excluding A and D | -0.140 [-0.145,-0.135] |
|  | MI including A and D | -0.146 [-0.150,-0.141] |
|  | NARFCS excluding A and D | 0.00250 [0.00129, 0.00370] |
|  | NARFCS including A and D | 0.00255 [0.00136, 0.00375] |

a: Missing Completely At Random; b: Missing At Random; c: Missing Not At Random.

#### **3.3 Evaluation of IPW in the simplified simulation study setting**

We evaluated inverse probability weighting assuming MAR (IPW), using unstabilised and stabilised weights, in the simpler simulation study where only outcome $Y$ was partially observed (as described in supplementary section 3.2). Briefly we considered three missingness mechanisms $Y$ MCAR, $Y$ MAR given $X,A_{1},A_{2},$ and $D$, and $Y$ MNAR given $Y,X,A_{1},A_{2},$ and $D$.

Since all covariates and auxiliary variables were fully observed, we only considered the following weighting model

$Pr\left( R^{Y}=1 \right)=expit\left\{ \rho_{0}+\rho_{X}X+\rho_{A_{1}}A_{1}+\rho_{A_{2}}A_{2}+\rho_{D}D \right\}$,

where $R^{Y}=1$ when $Y$ was observed and $R^{Y}=0$ otherwise, which was fitted to all participants. The unstabilised and stabilised weights were generated as described in supplementary section 1.6.

Supplementary tables 13 and 14 show the CCA and IPW results for $\beta_{X}$ when the true value of $\beta_{X}$ was $\ln3$ and $0$, respectively. As expected, for $Y$ MCAR the CCA and IPW results were unbiased with nominal CI coverage. For $Y$ MAR and $\beta_{X}=\ln3$ the CCA results were biased with poor CI coverage but applying IPW with a correctly specified weighting model eliminated most of the bias due to missing data (with vastly improved CI coverage). Note that for $\beta_{X}=0$ the CCA estimate was unbiased because in this scenario missingness was independent of $Y$ after conditioning on $X,W,$ and $Z$ (supplementary figure 1). Hence IPW was not required to remove bias in this scenario. For both the $\beta_{X}=\ln3$ and $\beta_{X}=0$ scenarios, the empirical standard error of IPW with unstabilised weights was more than double that of CCA due to some large weights. Also, we note that this empirical standard error was substantially larger for the $\beta_{X}=0$ scenario than the $\beta_{X}=\ln3$ scenario. A likely explanation is that for the $\beta_{X}=0$ scenario the auxiliary variables ($A_{1},A_{2},$ and $D$) only predicted $R^{Y}$ (i.e., did not predict the values of $Y$) and IPW would still be valid if these auxiliary variables were omitted from the weighting model. In this situation, including these auxiliary variables in the weighting model can lead to an increase in the variability of the weights (4). Applying IPW with stabilised weights greatly reduced the empirical standard error, with virtually unbiased estimates and nominal CI coverage.

As expected for $Y$ MNAR, all estimates were biased with poor CI undercoverage.

Note that for $\beta_{X}=\ln3$, the level of bias of the CCA estimate of $\beta_{X}$ was designed to be similar for the MAR and MNAR mechanisms. We achieved this by setting the auxiliary variables to be stronger predictors of missingness in the MAR mechanism compared to the MNAR mechanism. For $\beta_{X}=\ln3$, using stabilised weights reduced the empirical standard error but also resulted in an increase in the level of bias (compared to IPW with unstabilised weights) because lower variability in the estimates of $\beta_{X}$ led to fewer over-estimates compensating for the negative bias induced by the missing data. Due to the absence of large weights when $\beta_{X}=0$, the results of IPW with unstabilised and stabilised weights were very similar.

**Supplementary table 13:** Summary of the results for complete case analysis (CCA) and inverse probability weighting assuming missing at random (IPW) from the simplified simulation study when $\beta_{X}=\ln3$: Bias and empirical standard error (Emp SE) of exposure effect estimate $\hat{\beta}_{X}$, mean of the model-based standard error (SE) of $\hat{\beta}_{X}$, empirical coverage percentage of the 95% confidence interval for $\hat{\beta}_{X}$. [95% Monte Carlo interval].

| **Missingness mechanism of** $\boldsymbol{Y}$ | **Missingness method** | **Bias** | **Emp. SD** | **Mean SE^$^** | **Coverage %** |
| --- | --- | --- | --- | --- | --- |
| MCAR^$^ | CCA | 0.00164 [-0.00292, 0.00620] | 0.0736 [0.0704, 0.0769] | 0.0740 [0.0738, 0.0742] | 94.8 [93.4, 96.2] |
|  | IPW; unstabilised weights | 0.00162 [-0.00294, 0.00618] | 0.0735 [0.0703, 0.0768] | 0.0739 [0.0737, 0.0741] | 94.7 [93.3, 96.1] |
|  | IPW; stabilised weights | 0.00168 [-0.00288, 0.00624] | 0.0735 [0.0703, 0.0768] | 0.0739 [0.0737, 0.0741] | 94.8 [93.4, 96.2] |
| MAR^&^ | CCA | -0.130 [-0.133, -0.128] | 0.0441 [0.0421, 0.0461] | 0.0443 [0.0443,.0444] | 17.5 [15.1, 19.9] |
|  | IPW; unstabilised weights | -0.0152 [-0.00922, -0.0211] | 0.0960 [0.0918, 0.100] | 0.0925 [0.0900, 0.0949] | 90.2 [88.4, 92.0] |
|  | IPW; stabilised weights | 0.00807 [0.00384, 0.0123] | 0.0683 [0.0653, 0.0713] | 0.0671 [0.0669, 0.0674] | 94.6 [93.2, 96.0] |
| MNAR^*^ | CCA | -0.136 [-0.140, -0.132] | 0.0675 [0.0646, 0.0705] | 0.0648 [0.0647, 0.0649] | 44.0 [40.9, 47.1] |
|  | IPW; unstabilised weights | -0.140 [-0.148, -0.131] | 0.137 [0.131, 0.143] | 0.128 [0.127, 0.129] | 78.4 [75.8, 81.0] |
|  | IPW; stabilised weights | -0.150 [-0.156, -0.145] | 0.0878 [0.0840, 0.0917] | 0.0863 [0.0859, .0867] | 57.3 [54.2, 60.4] |

$ MCAR: Missing completely at random; & MAR: Missing at random; * MNAR: Missing not at random

**Supplementary table 14:** Summary of the results for complete case analysis (CCA) and inverse probability weighting assuming missing at random (IPW) from the simplified simulation study when $\beta_{X}=0$: Bias and empirical standard error (Emp SE) of exposure effect estimate $\hat{\beta}_{X}$, mean of the model-based standard error (SE) of $\hat{\beta}_{X}$, empirical coverage percentage of the 95% confidence interval for $\hat{\beta}_{X}$. [95% Monte Carlo interval].

| **Missingness mechanism of** $\boldsymbol{Y}$ | **Missingness method** | **Bias** | **Emp. SD** | **Mean SE^$^** | **Coverage %** |
| --- | --- | --- | --- | --- | --- |
| MCAR^$^ | CCA | 0.000469 [-0.00358, 0.00452] | 0.0654 [0.0625, 0.0682] | 0.0652 [0.0651, 0.0654] | 94.9 [93.5, 96.3] |
|  | IPW; unstabilised weights | 0.000544 [-0.00351, 0.00460] | 0.0654 [0.0625, 0.0682] | 0.0652 [0.0650, 0.0654] | 94.9 [93.5, 96.3] |
|  | IPW; stabilised weights | 0.000518 [-0.00353, 0.00457] | 0.0654 [0.0625, 0.0682] | 0.0652 [0.0650, 0.0654] | 95.0 [93.6, 96.4] |
| MAR^&^ | CCA | 0.00233 [-0.00155, 0.00621] | 0.0627 [0.0599, 0.0654] | 0.0655 [0.0654, 0.0657] | 96.5 [95.3, 97.6] |
|  | IPW; unstabilised weights | 0.0170 [0.00759, 0.0265] | 0.152 [0.146, 0.159] | 0.139 [0.137, 0.142] | 87.0 [84.9, 89.1] |
|  | IPW; stabilised weights | -0.0000977 [-.00577, .00557] | 0.0915 [0.0875, 0.0955] | 0.0940 [0.0935, 0.0946] | 95.3 [94.0, 96.6] |
| MNAR^*^ | CCA | -0.144 [-0.147, -0.140] | 0.0512 [0.0489, 0.0534] | 0.0508 [0.0507, 0.0509] | 19.4 [16.9, 21.9] |
|  | IPW; unstabilised weights | -0.134 [-0.137, -0.130] | 0.0519 [0.0496, 0.0542] | 0.0519 [0.0518, 0.0520] | 27.2 [24.4, 30.0] |
|  | IPW; stabilised weights | -0.135 [-0.138, -0.132] | 0.0517 [0.0494, 0.0539] | 0.0517 [0.0516, 0.0519] | 26.1 [23.4, 28.8] |

$ MCAR: Missing completely at random; & MAR: Missing at random; * MNAR: Missing not at random

#### **3.4 Additional simulation results for Monte Carlo NARFCS**

**Supplementary table 15:** Summary of the results when applying Monte Carlo NARFCS with differing number of Monte Carlo steps and imputations when $\beta_{X}=\ln3$: Bias and empirical standard error (Emp SE) of estimate $\hat{\beta}_{X}$, mean of the model-based SE of $\hat{\beta}_{X}$, coverage of the 95% confidence interval for $\hat{\beta}_{X}$. [95% Monte Carlo interval]. Simulation using selection model data generating model. Results based on 1000 simulated datasets.

|  | **Bias** | **Emp. SE** | **Mean SE^$^** | **Coverage %** |
| --- | --- | --- | --- | --- |
| Very informative prior; 10000 Monte Carlo steps; 1 imputation | -0.00121 [-0.00240, -0.0000119] | 0.0193 [0.0184, 0.0201] | 0.0226 [0.0225, 0.0227] | 95.9 [94.7, 97.1] |
| Very informative prior; 10000 Monte Carlo steps; 5 imputations | -0.00113 [-0.00233, 0.0000588] | 0.0192 [0.0184, 0.0201] | 0.0240 [0.0239, 0.0241] | 97.1 [96.1, 98.1] |
| Very informative prior; 5000 Monte Carlo steps; 1 imputation | -0.00123 [-0.00242, -0.0000347] | 0.0192 [0.0184, 0.0201] | 0.0226 [0.0225, 0.0227] | 95.9 [94.7, 97.1] |
| Informative prior; 10000 Monte Carlo steps; 1 imputation | -0.00358 [-0.00479, -0.00237] | 0.0195 [0.0186, 0.0203] | 0.0385 [0.0380, 0.0391] | 97.6 [96.7, 98.5] |
| Informative prior; 10000 Monte Carlo steps; 5 imputations | -0.00343  [-0.00464, -0.00223] | 0.0194 [0.0186, 0.0203] | 0.0408 [0.0402, 0.0413] | 98.1 [97.3, 98.9] |
| Informative prior; 5000 Monte Carlo steps; 1 imputation | -0.00354  [-0.00475, -0.00234] | 0.0195  [0.0186, 0.203] | 0.0386  [0.0380, 0.0391] | 97.6  [96.7, 98.5] |
| Vague prior; 10000 Monte Carlo steps; 1 imputation | -0.215 [-0.219, -0.211] | 0.0678 [0.0648, 0.0707] | 0.489 [0.488, 0.490] | 94.4 [93.0, 95.8] |
| Vague prior; 10000 Monte Carlo steps; 5 imputations | -0.216 [-0.221, -0.212] | 0.0677 [0.0647, 0.0706] | 0.490 [0.489, 0.491] | 95.1 [93.8, 96.4] |
| Vague prior; 5000 Monte Carlo steps; 1 imputation | -0.215  [-0.219, -0.210] | 0.0680  [0.0650, 0.0709] | 0.489  [0.488, 0.490] | 94.9  [93.5, 96.3] |

$ Standard deviation of the frequency distribution of $\hat{\beta}_{X}^{\delta^{NARFCS(1)}},\hat{\beta}_{X}^{\delta^{NARFCS(2)}},\cdots,\hat{\beta}_{X}^{\delta^{NARFCS(S)}}$.

**Supplementary table 16:** Summary of the results when applying Monte Carlo NARFCS with differing number of Monte Carlo steps and imputations when $\beta_{X}=0$: Bias and empirical standard error (Emp SE) of estimate $\hat{\beta}_{X}$, mean of the model-based SE of $\hat{\beta}_{X}$, coverage of the 95% confidence interval for $\hat{\beta}_{X}$. [95% Monte Carlo interval]. Simulation using selection model data generating model. Results based on 1000 simulated datasets.

|  | **Bias** | **Emp. SE** | **Mean SE^$^** | **Coverage %** |
| --- | --- | --- | --- | --- |
| Very informative prior; 10000 Monte Carlo steps; 1 imputation | -0.00206 [-0.00314, -0.000975] | 0.0175 [0.0167, 0.0182] | 0.0231  [0.0229, 0.0233] | 97.3 [96.3, 98.3] |
| Very informative prior; 10000 Monte Carlo steps; 5 imputations | -0.00192 [-0.00300, -0.000840] | 0.0175 [0.0167, 0.0182] | 0.0242 [0.0240, 0.0244] | 97.7 [96.8, 98.6] |
| Very informative prior; 5000 Monte Carlo steps; 1 imputation | -0.00209  [-0.00317, -0.00101] | 0.0175  [0.0167, 0.0182] | 0.0231  [0.0229, 0.0233] | 97.3  [96.3, 98.3] |
| Informative prior; 10000 Monte Carlo steps; 1 imputation | -0.00379 [-0.00488, -0.00271] | 0.0175 [0.0168, 0.0183] | 0.0372  [0.0366, 0.0377] | 98.0 [97.1, 98.9] |
| Informative prior; 10000 Monte Carlo steps; 5 imputations | -0.00351 [-0.00459, -0.00242] | 0.0175 [0.0167, 0.0183] | 0.0387 [0.0382, 0.0393] | 98.5 [97.7, 99.3] |
| Informative prior; 5000 Monte Carlo steps; 1 imputation | -0.00380  [-0.00489, -0.00271] | 0.0175  [0.0168, 0.0183] | 0.0372  [0.0366, 0.0377] | 98.2  [97.4, 99.0] |
| Vague prior; 10000 Monte Carlo steps; 1 imputation | -0.106 [-0.109, -0.103] | 0.0470 [0.0449, 0.0490] | 0.0822  [0.0811, 0.0832] | 96.9 [95.8, 98.0] |
| Vague prior; 10000 Monte Carlo steps; 5 imputations | -0.105  [-0.108, -0.102] | 0.0468 [00447, 0.0488] | 0.0843 [0.0832, 0.0853] | 97.5 [96.5, 98.5] |
| Vague prior; 5000 Monte Carlo steps; 1 imputation | -0.106  [-0.109, -0.103] | 0.0470  [0.0449, 0.0491] | 0.0822  [0.0811, 0.0933] | 96.6  [95.5, 97.7] |

$ Standard deviation of the frequency distribution of $\hat{\beta}_{X}^{\delta^{NARFCS(1)}},\hat{\beta}_{X}^{\delta^{NARFCS(2)}},\cdots,\hat{\beta}_{X}^{\delta^{NARFCS(S)}}$.

#### **3.5 Additional simulation results for Bayesian SM**

##### **3.5.1 Results not returned by Bayesian SM**

The Bayesian SM method did not return results for <10% of the simulated datasets. There were two causes of failure: (1) initial values for the model parameters of Bayesian SM. The method returned results when we re-ran the model with different initial values. The initial values were chosen randomly by sampling from a Normal distribution with mean 0 and variance 4. Occasionally, the combination of initial values caused the method to fail. (2) Non-positive definite matrix for the variance-covariance matrix of the linear regression for $X$. This second problem occurred for all three priors for $\delta^{SM}$ and when using different initial values.

Supplementary table 17 shows the number of failed results according to the four simulation scenarios and three priors for $\delta^{SM}$. The slightly lower number of failed results (due to non-positive definite variance-covariance matrix) for SM data generating model compared to PMM data generating model was likely because the bias model of Bayesian SM more closely resembled the SM data generating model.

**Supplementary table 17**: Number of simulated datasets (% out of 1000) where Bayesian SM failed to return results according to cause of failure.

| **Data generating model** | **True value of** $\boldsymbol{\beta}_{\boldsymbol{X}}$ | **Prior for** $\boldsymbol{\delta}^{\boldsymbol{SM}}$ | **Poor starting values** | **Non-positive covariance matrix** |
| --- | --- | --- | --- | --- |
| Selection model | $\ln3$ | very informative | 1 (0.2%) | 72 (7.2%) |
|  |  | informative | 0 (0%) | 72 (7.2%) |
|  |  | vague | 2 (0.2%) | 72 (7.2%) |
| Selection model | $0$ | very informative | 1 (0.1%) | 71 (7.1%) |
|  |  | informative | 1 (0.1%) | 71 (7.1%) |
|  |  | vague | 0 (0%) | 71 (7.1%) |
| Pattern-mixture model | $\ln3$ | very informative | 0 (0%) | 79 (7.9%) |
|  |  | informative | 1 (0.1%) | 79 (7.9%) |
|  |  | vague | 1 (0.1%) | 78 (7.8%) |
| Pattern-mixture model | $0$ | very informative | 0 (0%) | 84 (8.4%) |
|  |  | informative | 1 (0.1%) | 84 (8.4%) |
|  |  | vague | 2 (0.2%) | 84 (8.4%) |

##### **3.5.2 Convergence of Bayesian SM**

In this section we describe our investigations into why the bias and empirical standard error of the Bayesian SM estimate of $\beta^{X}$ were larger than expected.

Let $\hat{\beta}_{X}-truth$ denote the deviation of the estimate of $\beta_{X}$ from its true value. Supplementary figure 2 shows histograms of the deviations of the Bayesian SM estimates across the simulated datasets for data generated using SM data generating model. These deviations tended to be left-skewed, and the degree of skewness was greater when the true exposure effect was $\ln3$ and when the prior distribution for bias parameter $\delta^{SM}$ was vague.

The histograms show that there were some outlying values among the deviations of the Bayesian SM estimates. We suspected that these outliers were due to nonconvergence of the JAGS sampler. To investigate, we re-ran the Bayesian SM analysis with multiple chains in a random sample of those datasets corresponding to the outliers in supplementary figure 2. Running Bayesian SM with multiple chains resulted in estimates of $\beta_{X}$ closer to the truth.

The nonconvergence issue occurred because we analysed each simulated dataset with the same number of burn-in iterations. Given the time it took to run the model on a single dataset,

it was not possible to set the chain length to a value that would guarantee convergence for every simulated dataset. In practice when analysing a real dataset, non-convergence would be identified at the diagnostic stage and an appropriate burn-in length would be specified.

|  | $\boldsymbol{\beta}_{\boldsymbol{X}}\boldsymbol{=}\ln\boldsymbol{3}$ | $\boldsymbol{\beta}_{\boldsymbol{X}}\boldsymbol{=}\boldsymbol{0}$ |
| --- | --- | --- |
| **Very**    **Informative** | 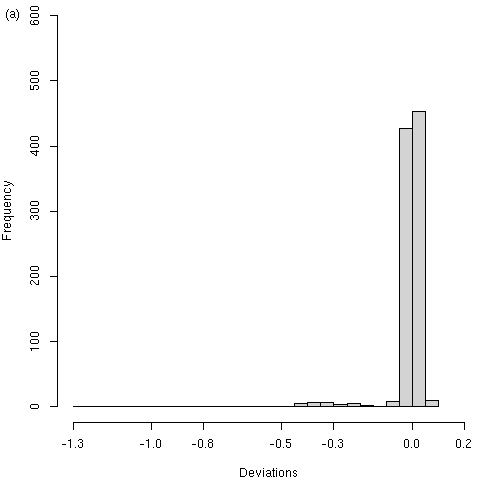 | 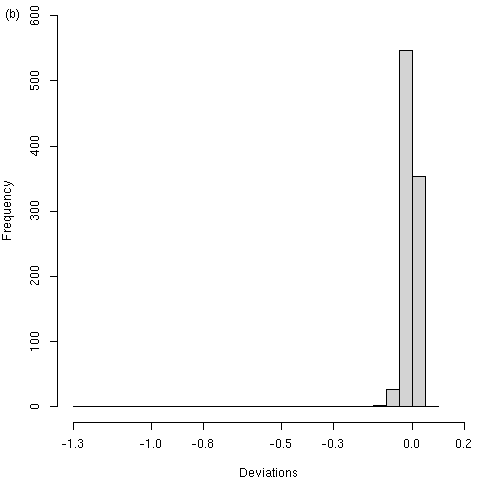 |
| **Informative** | 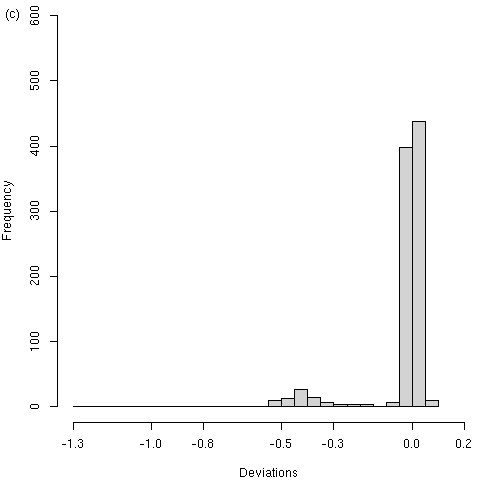 | 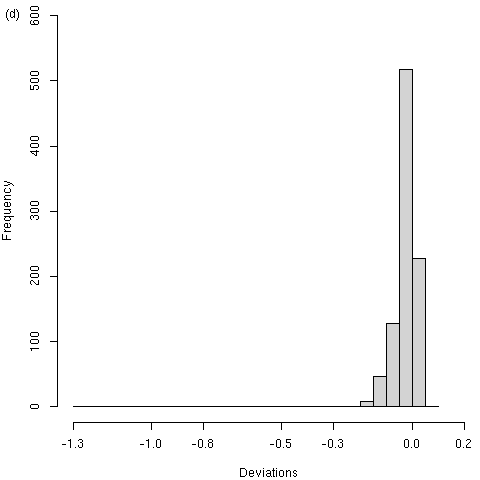 |
| **Vague** | 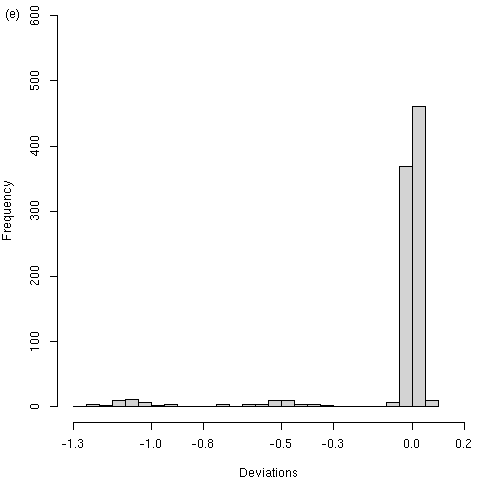 | 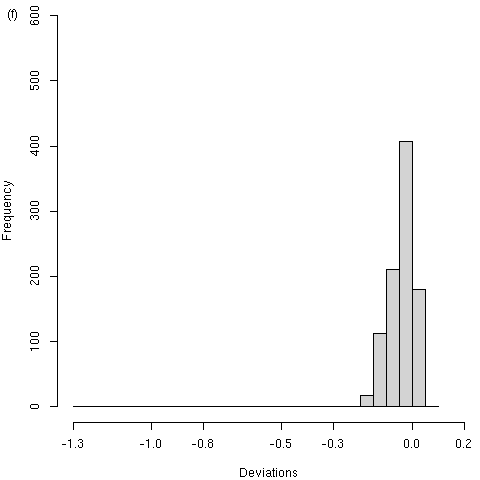 |
| **Supplementary Figure 2:** Histograms of deviations of the Bayesian SM exposure effect estimate from its true value (i.e., $\hat{\beta_{X}}-truth$) across datasets simulated under SM data generating model, according to $\beta_{X}=\ln3$ [left-hand column (a), (c), (e)] and $\beta_{X}=0$ [right-hand column (b), (d), (f)], and prior for $\delta^{SM}$: very informative [top row (a), (b)], informative [middle row (c), (d)] vague [bottom row (e), (f)]. | | |

#### **3.6 Main results for pattern-mixture model data generating model**

**Supplementary table 18:** Summary of results when $\beta_{X}=\ln3$ and simulation using pattern-mixture data generating model: number of simulated datasets (No. Sim.), bias and empirical standard error (Emp SE) of estimate $\hat{\beta}_{X}$, mean of model-based SE of $\hat{\beta}_{X}$, coverage of 95% confidence interval for $\hat{\beta}_{X}$. [95% Monte Carlo interval].

|  | **No.**  **Sim.** | **Bias** | **Emp. SD** | **Mean SE^$^** | **Coverage %** |
| --- | --- | --- | --- | --- | --- |
| Full data analysis | 1000 | -0.00138  [-0.00237, -0.000392] | 0.0160  [0.0153, 0.0167] | 0.0164  [0.0164, 0.0164] | 94.8  [93.4, 96.2] |
| Complete case analysis | 1000 | -0.162  [-0.166, -0.157] | 0.0724  [0.0692, 0.0755] | 0.0712  [0.0710, 0.0713] | 38.6  [35.6, 41.6] |
| MI | 1000 | -0.233 [-0.238, -0.229] | 0.0688 [0.0658, 0.0718] | 0.0681 [0.0677, 0.0686] | 7.40 [5.78, 9.02] |
| IPW; main model; unstabilised weights | 1000 | -0.214 [-0.223, -0.205] | 0.148 [0.142, 0.155] | 0.136 [0.135, 0.137] | 65.1 [62.1, 68.1] |
| IPW; main model; stabilised weights | 1000 | -0.179  [-0.185, -0.173] | 0.0986  [0.0943, 0.1029] | 0.0957 [0.0953, 0.0962] | 52.9 [49.8, 56.0] |
| Monte Carlo NARFCS^%^ with very informative prior | 1000 | -0.00400 [-0.00517, -0.00282] | 0.0190 [0.0181, 0.0198] | 0.0229 [0.0228, 0.0230] | 97.6 [96.7, 98.5] |
| Monte Carlo NARFCS^%^ with informative prior | 1000 | -0.00632 [-0.00751, -0.00513] | 0.0192 [0.0183, 0.0200] | 0.0395 [0.0389, 0.0400] | 100 [100, 100] |
| Monte Carlo NARFCS^%^ with vague prior | 1000 | -0.224 [-0.228, -0.220] | 0.0681 [0.0651, 0.0711] | 0.0514 [0.0512, 0.0515] | 100 [100, 100] |
| Bayesian SM with very informative prior | 921 | -0.0125  [-0.0143, -0.0107] | 0.0280  [0.0267, 0.0292] | 0.0261 [0.0237, 0.0285] | 94.1  [92.6, 95.7] |
| Bayesian SM with informative prior | 920 | -0.0291  [-0.0351, -0.0231] | 0.0929  [0.0887, 0.0972] | 0.0315 [0.0288, 0.0342] | 90.9  [89.0, 92.7] |
| Bayesian SM with vague | 921 | -0.0506  [-0.0621, -0.0392] | 0.177  [0.169, 0.185] | 0.0546 [0.0475, 0.0616] | 89.5  [87.5, 91.5] |

$ For Bayesian SM: standard deviation of the posterior distribution of $\hat{\beta}_{X}$, and for Monte Carlo NARFCS: standard deviation of the frequency distribution of $\hat{\beta}_{X}$. % Monte Carlo NARFCS using 10,000 Monte Carlo steps with single imputation.

**Supplementary table 19:** Summary of results when $\beta_{X}=0$ and simulation using pattern-mixture data generating model: number of simulated datasets (No. Sim.), bias and empirical standard error (Emp SE) of estimate $\hat{\beta}_{X}$, mean of model-based SE of $\hat{\beta}_{X}$, coverage of 95% confidence interval for $\hat{\beta}_{X}$. [95% Monte Carlo interval].

|  | **No.**  **Sim.** | **Bias** | **Emp. SD** | **Mean SE^$^** | **Coverage %** |
| --- | --- | --- | --- | --- | --- |
| Full data analysis | 1000 | 0.000415  [-0.000483, 0.00131] | 0.0145  [0.0138, 0.0151] | 0.0145  [0.0145, 0.0146] | 96.0  [94.8, 97.2] |
| Complete case analysis | 1000 | -0.134  [-0.138, -0.131] | 0.0546  [0.0522, 0.0570] | 0.0542  [0.0541, 0.0543] | 29.6  [26.8, 32.4] |
| MI | 1000 | -0.128 [-0.131, -0.125] | 0.0521 [0.0498, 0.0544] | 0.0524 [0.0521, 0.0528] | 31.1 [28.2, 34.0] |
| IPW; main model; unstabilised weights | 1000 | -0.126 [-0.129, -0.122] | 0.0570 [0.0545, 0.0595] | 0.0566 [0.0565, 0.0568] | 39.6 [36.6, 42.6] |
| IPW; main model; stabilised weights | 1000 | -0.127 [-0.130, -0.123] | 0.0570 [0.0545, 0.0595] | 0.0565 [0.0563, 0.0566] | 39.1 [36.1, 42.1] |
| Monte Carlo NARFCS^%^ with very informative prior | 1000 | -0.00135 [-0.00239, -0.000309] | 0.0168 [0.0160, 0.0175] | 0.0229 [0.0227, 0.0231] | 98.5 [97.7, 99.3] |
| Monte Carlo NARFCS^%^ with informative prior | 1000 | -0.00307 [-0.00412, -0.00203] | 0.0169 [0.0161, 0.0176] | 0.0367 [0.0362, 0.0373] | 99.7 [99.4, 100] |
| Monte Carlo NARFCS^%^ with vague prior | 1000 | -0.101 [-0.104, -0.0983] | 0.0472 [0.0451, 0.0493] | 0.0806 [0.0795, 0.0816] | 99.5 [99.1, 99.9] |
| Bayesian SM with very informative prior | 916 | -0.00730  [-0.00869, -0.00592] | 0.0214  [0.0204, 0.0224] | 0.0348 [0.0340, 0.0357] | 98.9  [98.2, 99.6] |
| Bayesian SM with informative prior | 915 | -0.0306  [-0.0333, -0.0280] | 0.0406  [0.0387, 0.0424] | 0.0443 [0.0435, 0.0451] | 90.6  [88.7, 92.5] |
| Bayesian SM with vague | 914 | -0.0489  [-0.0522, -0.0457] | 0.0502  [0.0479, 0.0525] | 0.0459 [0.0451, 0.0467] | 78.9  [76.2, 81.5] |

$ For Bayesian SM: standard deviation of the posterior distribution of $\hat{\beta}_{X}$, and for Monte Carlo NARFCS: standard deviation of the frequency distribution of $\hat{\beta}_{X}$. % Monte Carlo NARFCS using 10,000 Monte Carlo steps with single imputation.

### **4 References**

| 1. | Tompsett DM, Leacy F, Moreno-Betancu M, Heron J, White IR. On the use of the not-at-random fully conditional specification (NARFCS) procedure in practice. Statistics in Medicine. 2018; 37: 2338-2353. |
| --- | --- |
| 2. | Ward H, Atchison C, Whitaker M, Ainslie KE, Elliott J, Okell L, et al. SARS-CoV-2 antibody prevalence in England following the first peak of the pandemic. Nature communications. 2021; 12(1): 905. |
| 3. | Thoemmes F, Rose N. A Cautious Note on Auxiliary Variables That Can Increase Bias in Missing Data Problems. Multivariate Behavioral Research. 2014; 49: 443-459. |
| 4. | Seaman SR, White IR. Review of inverse probability weighting for dealing with missing data. Statistical Methods in Medical Research. 2013; 22(3): 278-295. |
